# Time-to-event modeling with multimodal clinical and genetic features improves risk stratification of liver complications in chronic hepatitis C

**DOI:** 10.64898/2026.03.06.26347819

**Authors:** Humayera Islam, Ash Arian, Joseph W. Franses, Habibul Ahsan

## Abstract

Chronic hepatitis C (CHC) remains a leading cause of cirrhosis, hepatocellular carcinoma (HCC), and premature mortality despite effective antiviral therapy, underscoring the need for individualized risk stratification beyond fibrosis stage alone. Using harmonized data from the All of Us Research Program, we developed and internally validated an interpretable multimodal survival framework to predict incident cirrhosis, HCC, and all-cause mortality, explicitly accounting for competing death. Baseline predictors within a ±180-day window around CHC diagnosis included demographics, comorbidities, medications, laboratory biomarkers, socioeconomic context, and selected germline variants. Penalized Cox, ensemble, gradient-boosted, and neural survival models were compared under a consistent training and held-out testing strategy. Best-performing models achieved test C-indices of 0.67 for cirrhosis (Coxnet-LASSO), 0.71 for HCC, and 0.75 for mortality (Random Survival Forest), with stable time-dependent AUROC up to 0.81. Substantial feature compression preserved discrimination: restricting to the top 50% or 25% of predictors resulted in minimal absolute change in test performance (3.5%). Reduced models were anchored in clinically interpretable domains, including age, liver injury markers, hepatic reserve, cardiometabolic burden, deprivation index, and chromosome 19/22 loci. Feature importance reinforces existing known clinical and biological risk factors for liver complications: liver injury markers were most influential for cirrhosis and HCC, whereas hepatic reserve and cardiometabolic burden were more predictive of mortality, with age serving as a central baseline determinant across outcomes. Together, these results support a scalable and parsimonious framework for individualized CHC risk stratification that integrates multimodal determinants.

## Introduction

Chronic hepatitis C (CHC) affects an estimated 71 million individuals worldwide and remains a leading cause of cirrhosis, hepatocellular carcinoma (HCC), and liver-related mortality, accounting for nearly 290,000 deaths annually ^1^. Although direct-acting antivirals (DAAs) achieve sustained virologic response (SVR) rates exceeding 95%, long-term risk of cirrhosis, HCC, and premature death persists after viral clearance, particularly among individuals with advanced fibrosis, metabolic comorbidity, or delayed treatment access ^2–6^.

Current surveillance strategies are largely anchored to fibrosis stage. Guidelines recommend semiannual ultrasound screening for patients with cirrhosis (Fibrosis stage 4), while recommendations for advanced fibrosis (Fibrosis stage 3) remain variable ^5–7^. Risk refinement commonly relies on non-invasive fibrosis indices such as the Fibrosis-4 (FIB-4) index, AST to platelet ratio index (APRI), platelet count, albumin, bilirubin, and elastography ^8^. However, these measures are static and incompletely capture the heterogeneity of disease progression. Individuals within the same fibrosis category may experience markedly different trajectories, and competing mortality is rarely incorporated into routine risk estimation ^9^. Consequently, stage-based frameworks may misclassify risk and limit individualized surveillance strategies.

Progression of CHC reflects complex interactions among persistent immune activation, metabolic dysfunction, environmental exposures, and host susceptibility. Older age and longer untreated duration accelerate fibrosis advancement ^10–12^. Metabolic conditions such as diabetes and obesity increase risk of cirrhosis and HCC independent of viral status ^13,14^. Genetic variants (e.g., *PNPLA3*) and interferon-response loci contribute to inter-individual variability, while social determinants of health influence access to treatment access and long-term disease outcomes ^15–20^. Collectively, these findings underscore that progression risk extends well beyond fibrosis stage alone.

Survival modeling approaches have been applied to estimate CHC outcomes, including Cox proportional hazards and machine learning-based models ^8,21,22^. More recent landmark and competing-risk frameworks have improved dynamic prediction in selected cohorts ^23^. However, key limitations persist. Many models are derived from single-center or demographically homogeneous populations, limiting external validity ^7,24^. Competing mortality is not consistently incorporated, despite its clinical relevance ^9,25,26^. Integration of multimodal determinants, including clinical variables, socioeconomic factors, medication exposures and genetic variations, within large, diverse cohorts remains limited ^27–29^. Furthermore, few frameworks explicitly evaluate predictive performance with model parsimony, which limits clinical interpretability.

The All of Us (AoU) Research Program provides an opportunity to address these limitations. Designed to enroll over one million participants with intentional inclusion of populations historically underrepresented in biomedical research, AoU integrates longitudinal electronic health records (EHRs), physical measurements, survey-derived social determinants of health, and genomic sequencing within a harmonized cloud-based infrastructure ^30–32^. This multimodal framework enables development and evaluation of survival models in diverse, real-world CHC populations. Although challenges such as data heterogeneity, missingness, and temporal alignment require careful methodological handling ^32–34^, they also enable an opportunity to testing of robust and generalizable modeling strategies at scale.

In this study, we develop and internally validate an interpretable, multimodal survival modeling framework to predict incident cirrhosis, incident HCC, and all-cause mortality among individuals with CHC within the AoU cohort. Using the earliest documented CHC diagnosis as the time origin, we model disease progression and mortality within a framework that explicitly incorporates competing risks, recognizing that death alters the probability of observing subsequent liver-specific outcomes. Our aims are threefold: (1) to integrate harmonized EHR-derived clinical variables, laboratory biomarkers, medication exposures, socioeconomic indicators, and selected germline variants, and systematically compare penalized regression, ensemble machine learning, and neural survival approaches under a unified validation strategy, (2) to identify parsimonious feature sets that preserve predictive performance, while enhancing interpretability and implementation feasibility, and (3) to evaluate model discrimination across clinically relevant time horizons and examine robustness across racial, ethnic, and socioeconomic subgroups.

By leveraging multimodal data within a diverse national cohort, we propose an interpretable and scalable survival modeling framework that refines stage-based surveillance and enables individualized risk quantification of liver complications in chronic hepatitis C.

## Methods

### Data Source

This retrospective cohort study leveraged data from the AoU Research Program Controlled Tier Dataset (v8), accessed through the secure Researcher Workbench. Data in AoU are harmonized using the Observational Medical Outcomes Partnership (OMOP) Common Data Model, enabling standardized integration of heterogeneous data sources across participating healthcare systems.

Electronic health record (EHR) data comprise provider-derived information on clinical encounters, diagnoses, procedures, and medication exposures. Demographic characteristics and lifestyle information were self-reported by patients at enrollment through structured surveys. Mortality information was ascertained through linkage of EHR-derived death records and deceased status reporting within HealthPro. Clinical concepts were encoded using standardized vocabularies, including ICD-9/10, SNOMED-CT, and LOINC, ensuring semantic consistency and cross-site interoperability.

Genomic data were obtained from the AoU controlled-access genomic release, which includes genotype array data and short- and long-read whole-genome sequencing. These data capture single-nucleotide variants, insertions and deletions, and structural variants across a genetically diverse population and were linked to clinical records using participant identifiers within the secure Workbench environment.

All analyses complied with AoU data access policies, governance requirements, and ethical standards. The study was conducted under the AoU data use framework, and no individual-level data were exported outside the secure computing environment. Fig. 1 shows the modeling framework implemented in this study.

**Fig. 1.**
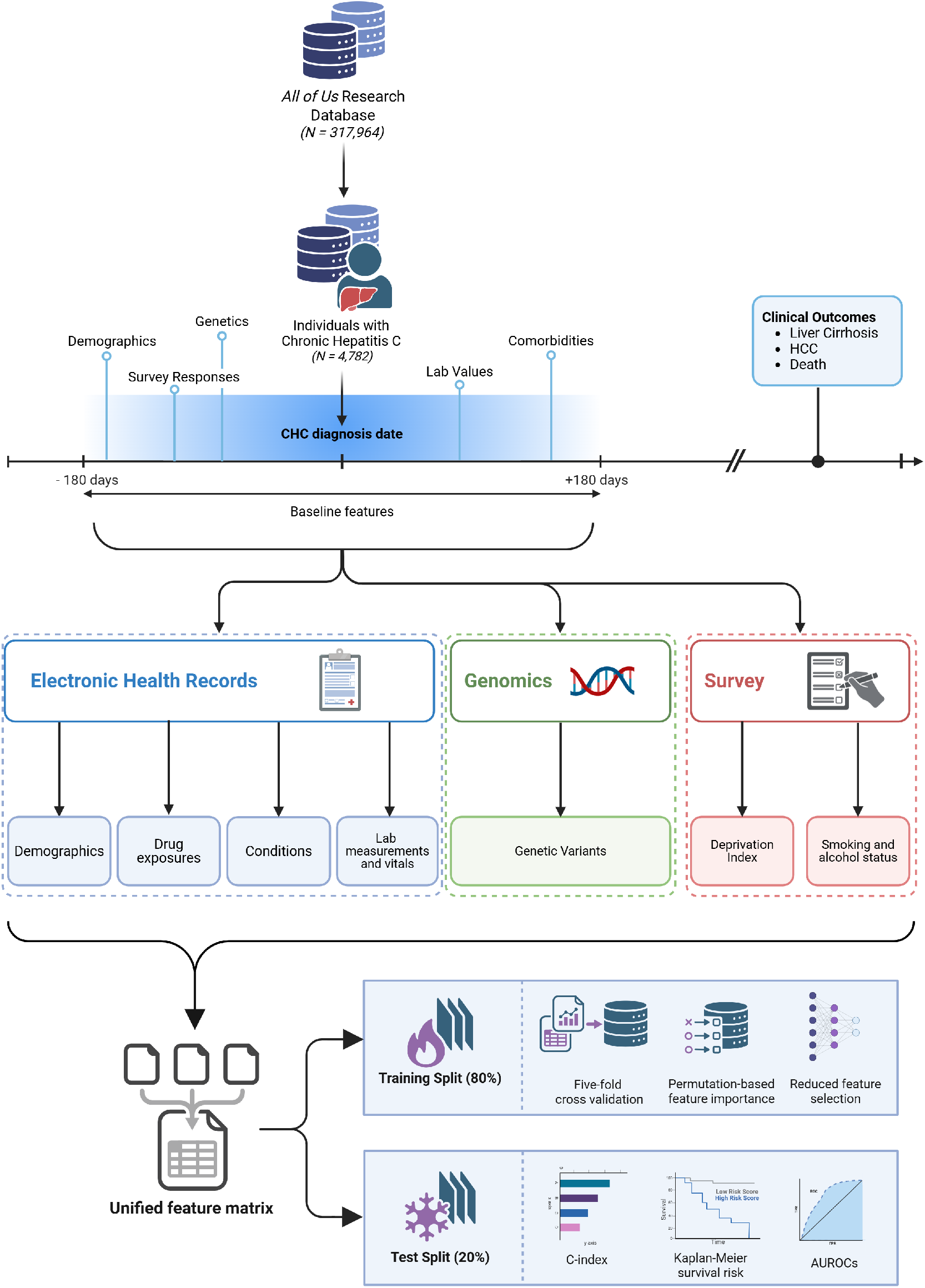
Survival modeling framework for chronic hepatitis C (CHC) outcomes. Individuals with CHC were identified from the All of Us Research Program, with the index date defined as the earliest documented CHC diagnosis. Baseline features were extracted within a ±180-day window surrounding the index date to preserve temporal validity and prevent outcome leakage. Multimodal predictors were derived from electronic health records, genomics, and survey data, including demographics, medication exposures, clinical conditions, laboratory measurements and vital signs, socioeconomic indicators such as deprivation index, lifestyle factors, and genetic variants. These variables were aggregated into a unified patient-level feature matrix. Time-to-event outcomes were defined as years from CHC diagnosis to incident liver cirrhosis, hepatocellular carcinoma (HCC), or all-cause mortality, with censoring at the last recorded encounter. The cohort was partitioned into training (80%) and held-out test (20%) sets. Within the training cohort, survival models were developed using five-fold cross-validation, permutation-based feature importance, and reduced feature selection (Top-25% and Top-50%). Model performance was evaluated in the held-out test set using Harrell’s concordance index (C-index), time-dependent AUROC, Kaplan–Meier risk stratification, SHAP-based interpretability, and subgroup analyses to ensure robust and clinically interpretable prognostic assessment.

### Chronic Hepatitis C Cohort

We identified a cohort of individuals with a documented diagnosis of chronic hepatitis C (CHC) using validated diagnosis concept sets mapped across ICD-9, ICD-10, SNOMED-CT, and OMOP concept identifiers (Supplementary Table 1). The index date was defined as the earliest recorded CHC diagnosis in the EHR. Particpants were required to be at least 18 years of age at baseline and to have longitudinal follow-up data available after diagnosis. To ensure reliable temporal characterization of disease onset, ±180-day window was applied around the index date. This window accounts for delays in diagnosis, documentation lag, and variability in care-seeking behavior. Individuals without any recorded clinical encounters within this window were excluded to reduce misclassification of disease onset and baseline features.

### Time-to-Event Outcomes

We evaluated three time-to-event outcomes: incident liver cirrhosis, incident HCC, and allcause mortality. Cirrhosis and HCC were identified using validated diagnosis concept sets corresponding to chronic liver cirrhosis and malignant liver neoplasm (Supplementary Table 1). Mortality status and dates were obtained from the AoU death table.

Time zero was defined as the earliest documented CHC diagnosis (index date). For cirrhosis and HCC, event time was defined as the interval from index date to the first occurrence of the respective diagnosis. For individuals without a recorded diagnosis of cirrhosis or HCC, death occurring prior to either liver-specific outcome was treated as a censoring event, with follow-up time calculated from the index date to the date of death ^9^. Patients who neither developed the outcome nor died were censored at their last documented clinical encounter within the EHR.

For all-cause mortality, event time was defined as the interval from index date to death, where death is not attributable to any specific cause, with right censoring at last documented follow-up. Event times and corresponding indicators were derived for each outcome and used in all survival modeling analyses.

### Survival Modeling Framework

To model progression from CHC diagnosis to each outcome of interest, we evaluated complementary survival modeling approaches, including classical statistical models, machine learning methods, and deep learning architectures.

Penalized Cox proportional hazards models were used as a baseline due to their interpretability and widespread clinical use. Both LASSO (L1) and Ridge regularization (L2) were evaluated to address high-dimensional covariates and multi-collinearity (LASSO emphasizes sparse feature selection and Ridge stabilizes correlated predictors).

To capture non-linear effects and potential departures from proportional hazards, we implemented Random Survival Forests (RSF), an ensemble method that extends random forests to right-censored data. By aggregating survival trees trained on bootstrap samples with log-rank splitting criteria, RSF can model complex feature-interactions and time-varying effects without explicit parametric assumptions. We also employed gradient-boosted survival models, which were trained on additive ensembles of weak learners optimized for survival objectives. Boosting enables sequential refinement of residual risk patterns and is effective in capturing subtle nonlinearities and interactions while maintaining strong predictive performance ^32^. In addition, a neural network-based Cox model (Cox-nnet v2) was trained to learn nonlinear feature representations while optimizing the Cox partial likelihood. The Cox-nnet v2 model includes one input layer, one hidden layer and one Cox regression output layer ^35^.

For each outcome, the cohort was randomly partitioned into a development set (80%) and an independent held-out test set (20%). All preprocessing steps were fit exclusively within the training data and subsequently applied to validation and test sets to prevent information leakage. Model development employed three-fold cross-validation within the training set for hyperparameter tuning and model selection. Final model performance was evaluated on the held-out test set.

### EHR Feature Engineering

We constructed a multimodal feature set encompassing demographic, clinical, medication, laboratory, social, and genetic variables. All features were extracted within a fixed ±180-day window centered on the CHC diagnosis index date to define baseline covariates (See details of feature extraction process in Supplementary Document Section 2).

Demographic and social features included age at CHC diagnosis, sex at birth, race, ethnicity, and an area-level deprivation index reflecting neighborhood socioeconomic context. Clinical comorbidities known to modify liver disease progression, including metabolic, cardiometabolic (e.g., hypertension, type 2 diabetes mellitus, gastroesophageal reflux disease), and substance use-related conditions, were encoded as binary indicators based on their documented presence within the feature window. Alcohol use and smoking status were derived from lifestyle survey data and treated as categorical variables to preserve clinically interpretable strata.

Medication exposures were represented as binary indicators reflecting whether an individual had evidence of exposure to major therapeutic classes (antibiotics, anticholesterol, anti-hypertensive, insulin, metformin use) within the ±180-day window. DAA therapy was modeled as a binary exposure indicating receipt versus non-receipt within this period, capturing early treatment initiation rather than cumulative dose or duration.

Laboratory biomarkers commonly used in liver (e.g., aspartate aminotransferase (AST), alanine aminotransferase (ALT), total bilirubin, and serum albumin), metabolic risk (e.g., hemoglobin A1c), and vitals (e.g., blood pressure) were summarized using mean values across all available measurements within the baseline window. Numerical features (e.g., labs, vitals, age, and deprivation index) were standardized by dividing by their root mean square, and variables exhibiting high skewness (absolute skewness >3) were log-transformed to stabilize distributions. Missing values in numerical features were handled using median imputation with explicit missingness indicators. Additionally, sensitivity to missingness was analyzed using other imputation techniques, including median imputation without indicators and K-nearest neighbors (KNN) imputation to assess robustness to imputation strategy.

Genomic features were constructed from a targeted panel of single nucleotide polymorphisms (SNPs) previously implicated in liver fibrosis progression, cirrhosis susceptibility, and HCC risk in genome-wide and integrative genetic studies ^18,19,36–40^. Selected loci included variants mapped to biologically relevant genes such as *SERPINA1* (chr14), *APOE/APOC1* (chr19), *TM6SF2* (chr19), *PNPLA3* (chr22), and additional loci associated with lipid metabolism, inflammation, and fibrogenesis. Genotype dosage data for predefined SNPs were extracted from the AoU genomic dataset and linked to the clinical cohort via participant identifiers. Genotypes were encoded as alternate allele counts (0, 1, or 2), corresponding to homozygous reference, heterozygous, and homozygous alternative, and modeled as categorical baseline covariates to preserve interpretability and avoid imposing linear additive assumptions. Patients with missing genomic features were excluded from the study. (Supplementary Table 2.6 and Supplementary Fig. 1).

### Reduced Feature Selection

To derive clinically interpretable models while preserving predictive performance, we employed a structured feature reduction strategy based on permutation importance scores ^35^. Permutation importance quantifies each feature’s marginal contribution to discrimination by measuring the change in performance following random permutation of that feature.

For each model and outcome, the permutation importance scores were computed within cross-validation using the three validation folds. Scores were averaged across folds to obtain stable feature rankings that reflect consistent predictive contribution rather than fold-specific variability. Importantly, permutation importance was computed on the trained full models, ensuring that feature rankings reflected performance in the presence of all available covariates.

Reduced models were then constructed using prespecified proportions of the ranked feature list, including the top 25% and top 50% of predictors. These reduced models were evaluated using the same training, cross-validation, and test framework as the full models, enabling direct comparison of discrimination and stability.

### Model Evaluation

The prediction targets were the time to the earliest occurrence of cirrhosis, HCC, and all-cause mortality from the index date of CHC diagnosis. Model performance was evaluated using Harrell’s concordance index (C-index), which assesses the ability of a model to correctly rank individuals according to their risk of experiencing an event over time. Cross-validation results are reported as the mean across folds, and test-set performance is reported with uncertainty quantified via nonparametric bootstrap resampling. Time-dependent area under the receiver operating characteristic curve (AUROC) was computed at 3-, 5-, and 10-year horizons to assess performance across follow-up time.

Kaplan–Meier survival curves were constructed to visualize differences in time-to-event distributions between patients stratified into low- and high-risk groups, and statistical separation was assessed using log-rank tests. Model explainability was assessed using SHapley Additive exPlanations (SHAP), enabling both global feature attribution and individual-level risk decomposition. To facilitate comparison with traditional statistical modeling approaches, Cox proportional hazards (CoxPH) models were fitted, and SHAP values were compared with CoxPH coefficients.

For cirrhosis and HCC outcomes, competing-risk analyses were additionally conducted to account for all-cause mortality. Cumulative incidence functions were estimated using the Aalen–Johansen estimator, and Fine–Gray subdistribution hazard models were fitted to estimate cause-specific risks. To characterize longitudinal disease trajectories, a nonparametric multi-state framework was implemented to model transitions from CHC to cirrhosis, HCC, and death, with state occupancy probabilities estimated using the Aalen–Johansen estimator.

## Results

### Cohort characteristics

The CHC cohort comprised 4,782 individuals, including 790 incident cirrhosis events, 91 incident HCC events, and 172 deaths during follow-up. The median age at CHC diagnosis was 51 years. The mean time from diagnosis to incident cirrhosis was 5.4 years, to HCC was 7.2 years, and to death was 7.4 years. Baseline demographic and clinical characteristics are summarized in Supplementary Table 8.

### Prediction of time to cirrhosis, HCC, and all-cause mortality

The complete set of analytic features comprised 42 variables, including demographic characteristics, age at CHC diagnosis, comorbidities, medication exposures, and selected genetic variants (complete list in Supplementary Table 7). Separate time-to-event models were developed for cirrhosis, HCC, and all-cause mortality. The cohort was split into 80% training data, evaluated using 3-fold cross-validation, and 20% as held-out testing data. Detailed characteristics of the training and test splits are provided in Supplementary Table 11.

Five survival modeling approaches were evaluated, and for each outcome, the model demonstrating the most consistent performance across experiments was selected as the final model (Table 1). During cross-validation, Harrell ’s C-index was 0.6710 ± 0.0224 for cirrhosis using Coxnet-LASSO, 0.7252 ± 0.0583 for HCC using Random Survival Forest (RSF), and 0.7595 ± 0.0463 for all-cause mortality using RSF. Discrimination remained consistent in the held-out test set, with C-indices of 0.6709 ± 0.0231 for cirrhosis, 0.7070 ± 0.0568 for HCC, and 0.7542 ± 0.0461 for mortality.

**Table 1:**
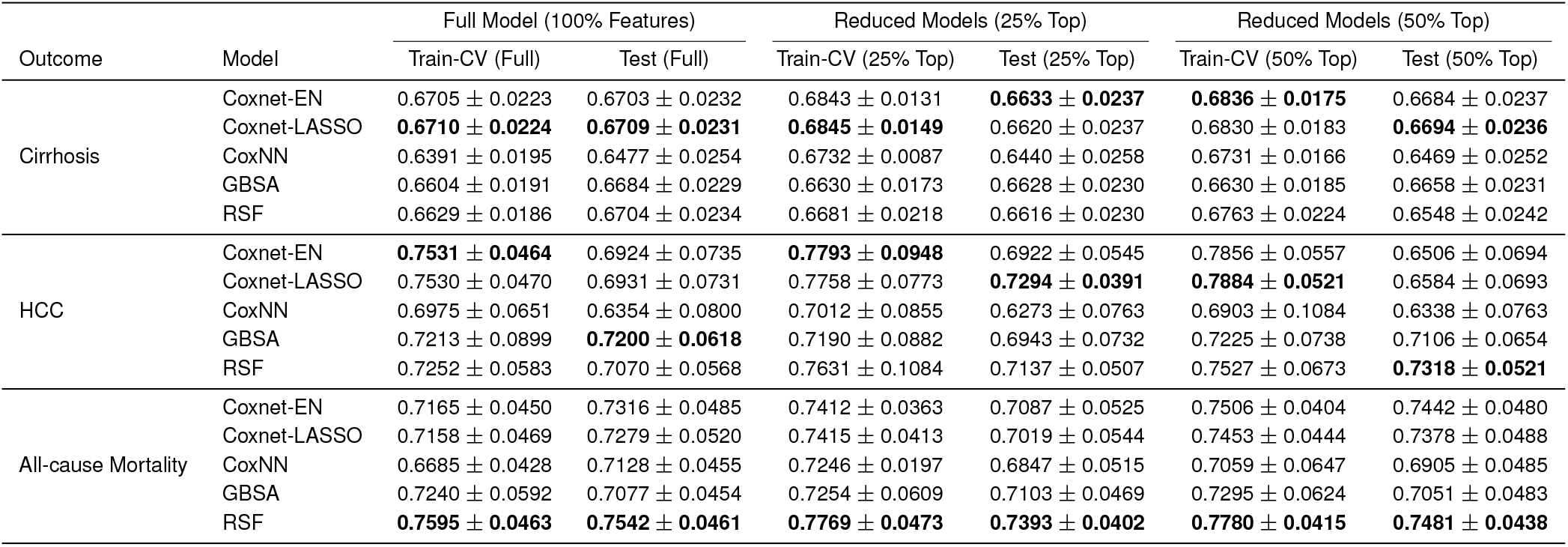
Discrimination performance (Harrell’s C-index, mean ±SD) of survival models for cirrhosis, hepatocellular carcinoma (HCC), and all-cause mortality using full and reduced feature sets. Performance is reported for five modeling approaches under 3-fold cross-validation in the training cohort and evaluation on a held-out test cohort. Reduced models retain either the top 25% or top 50% of features ranked by cross-validated permutation importance (ΔC-index).

| Outcome | Model | Full Model (100% Features) |  | Reduced Models (25% Top) |  | Reduced Models (50% Top) |  |
| --- | --- | --- | --- | --- | --- | --- | --- |
|  |  | Train-CV (Full) | Test (Full) | Train-CV (25% Top) | Test (25% Top) | Train-CV (50% Top) | Test (50% Top) |
| Cirrhosis | Coxnet-EN | 0.6705 ± 0.0223 | 0.6703 ± 0.0232 | 0.6843 ± 0.0131 | **0.6633 ± 0.0237** | **0.6836 ± 0.0175** | 0.6684 ± 0.0237 |
| | Coxnet-LASSO | **0.6710 ± 0.0224** | **0.6709 ± 0.0231** | **0.6845 ± 0.0149** | 0.6620 ± 0.0237 | 0.6830 ± 0.0183 | **0.6694 ± 0.0236** |
| | CoxNN | 0.6391 ± 0.0195 | 0.6477 ± 0.0254 | 0.6732 ± 0.0087 | 0.6440 ± 0.0258 | 0.6731 ± 0.0166 | 0.6469 ± 0.0252 |
| | GBSA | 0.6604 ± 0.0191 | 0.6684 ± 0.0229 | 0.6630 ± 0.0173 | 0.6628 ± 0.0230 | 0.6630 ± 0.0185 | 0.6658 ± 0.0231 |
| | RSF | 0.6629 ± 0.0186 | 0.6704 ± 0.0234 | 0.6681 ± 0.0218 | 0.6616 ± 0.0230 | 0.6763 ± 0.0224 | 0.6548 ± 0.0242 |
| HCC | Coxnet-EN | **0.7531 ± 0.0464** | 0.6924 ± 0.0735 | **0.7793 ± 0.0948** | 0.6922 ± 0.0545 | 0.7856 ± 0.0557 | 0.6506 ± 0.0694 |
| | Coxnet-LASSO | 0.7530 ± 0.0470 | 0.6931 ± 0.0731 | 0.7758 ± 0.0773 | **0.7294 ± 0.0391** | **0.7884 ± 0.0521** | 0.6584 ± 0.0693 |
| | CoxNN | 0.6975 ± 0.0651 | 0.6354 ± 0.0800 | 0.7012 ± 0.0855 | 0.6273 ± 0.0763 | 0.6903 ± 0.1084 | 0.6338 ± 0.0763 |
| | GBSA | 0.7213 ± 0.0899 | **0.7200 ± 0.0618** | 0.7190 ± 0.0882 | 0.6943 ± 0.0732 | 0.7225 ± 0.0738 | 0.7106 ± 0.0654 |
| | RSF | 0.7252 ± 0.0583 | 0.7070 ± 0.0568 | 0.7631 ± 0.1084 | 0.7137 ± 0.0507 | 0.7527 ± 0.0673 | **0.7318 ± 0.0521** |
| All-cause Mortality | Coxnet-EN | 0.7165 ± 0.0450 | 0.7316 ± 0.0485 | 0.7412 ± 0.0363 | 0.7087 ± 0.0525 | 0.7506 ± 0.0404 | 0.7442 ± 0.0480 |
| | Coxnet-LASSO | 0.7158 ± 0.0469 | 0.7279 ± 0.0520 | 0.7415 ± 0.0413 | 0.7019 ± 0.0544 | 0.7453 ± 0.0444 | 0.7378 ± 0.0488 |
| | CoxNN | 0.6685 ± 0.0428 | 0.7128 ± 0.0455 | 0.7246 ± 0.0197 | 0.6847 ± 0.0515 | 0.7059 ± 0.0647 | 0.6905 ± 0.0485 |
| | GBSA | 0.7240 ± 0.0592 | 0.7077 ± 0.0454 | 0.7254 ± 0.0609 | 0.7103 ± 0.0469 | 0.7295 ± 0.0624 | 0.7051 ± 0.0483 |
|  | RSF | **0.7595 ± 0.0463** | **0.7542 ± 0.0461** | **0.7769 ± 0.0473** | **0.7393 ± 0.0402** | **0.7780 ± 0.0415** | **0.7481 ± 0.0438** |

To enhance parsimony and clinical interpretability, feature reduction was performed using permutation importance rankings based on ΔC-index, as described in the Methods. The top 50% and 25% ranked features for each outcome are shown in Fig. 2(**d, e, f**), and comparative heatmaps of permutation importance across models are provided in Supplementary Fig. 2. Reduced models retaining the top 50% of features achieved performance comparable to that of the full models. For cirrhosis, the mean C-index was 0.6830 ± 0.0183 in cross-validation and 0.6694 ± 0.0236 in testing. For HCC, performance was 0.7631 ± 0.1084 (cross-validation) and 0.7318 ± 0.0521 (test). For mortality, the corresponding values were 0.7769 ± 0.0473 and 0.7481 ± 0.0438. Reducing to the top 25% of features, discrimination remained stable: cirrhosis 0.6681 ±0.0218 (cross-validation) and 0.6616 ± 0.0230 (test); HCC 0.7527 ± 0.0673 and 0.7137 ± 0.0507; mortality 0.7780 ±0.0415 and 0.7393 ± 0.0402. Comparisons between full and reduced models are shown in Fig. 2(**a, b, c**), with detailed results in Supplementary Tables 13, 15, and 17.

**Fig. 2.**
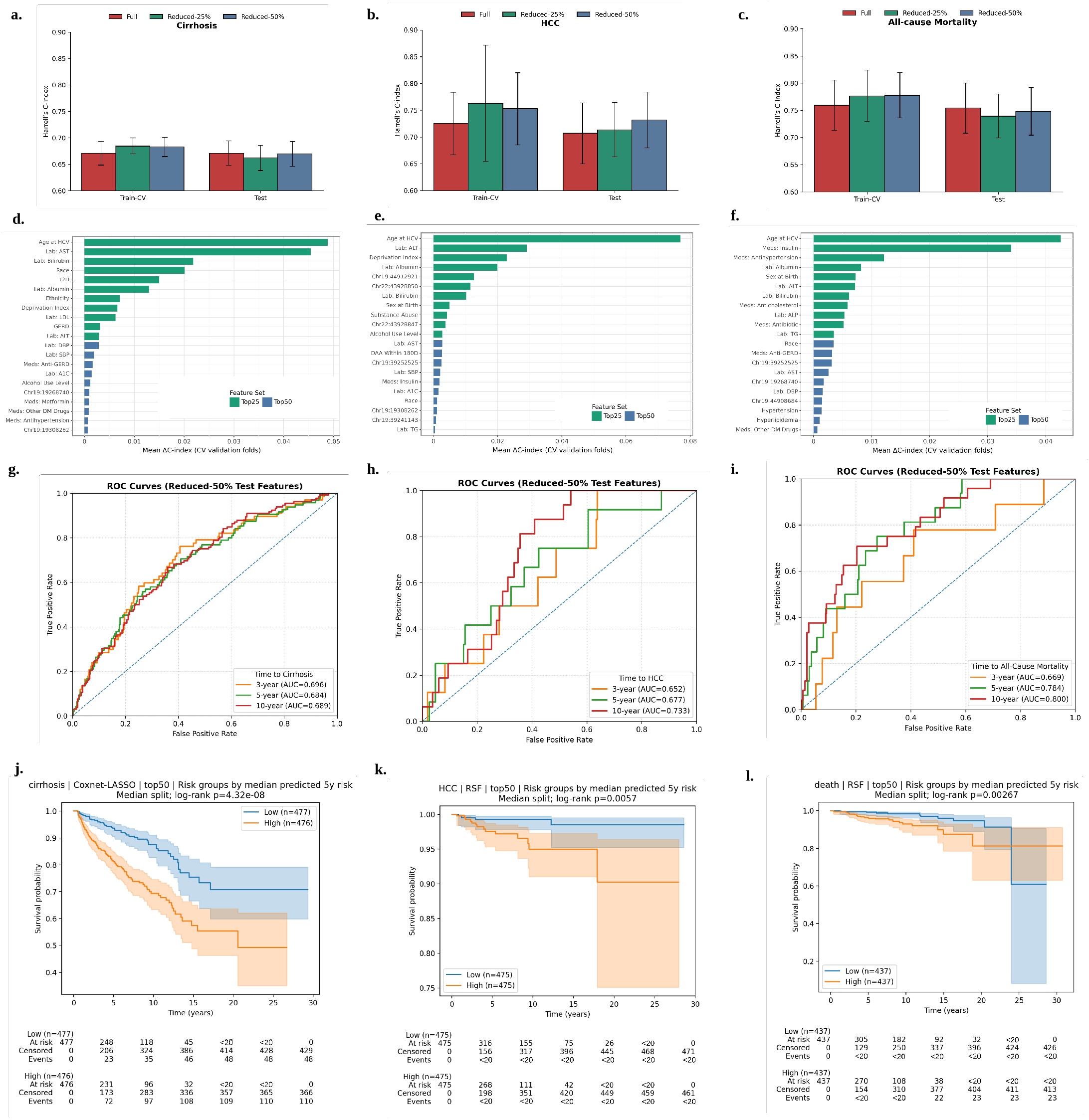
Model development, feature selection, and validation of reduced-risk prediction models for CHC-related outcomes.Overall performance and robustness. Panels **a, b**, and **c** summarize discrimination performance (Harrell’s C-index) for cirrhosis (Coxnet-LASSO), all-cause mortality (Random Survival Forest), and hepatocellular carcinoma (HCC; Random Survival Forest), respectively, using the best-performing model for each outcome. Performance is compared between full-feature models and reduced feature sets (top 25% and top 50%) under cross-validation and evaluation on a held-out test cohort. **Feature selection from training data**. Panels **d, e**, and **f** show permutation-based feature importance derived exclusively from cross-validation within the training cohort for cirrhosis (Coxnet-LASSO), all-cause mortality (RSF), and HCC (RSF), respectively. Features are ranked by mean Δ C-index across folds, and the top 50% feature sets selected from this stage are carried forward to downstream evaluation. **Time-dependent discrimination of reduced models**. Panels **g, h**, and **i** display time-dependent ROC curves at 3, 5, and 10 years for the reduced-50% models evaluated on the held-out test cohort, confirming sustained predictive performance over clinically relevant time horizons. **Risk stratification using reduced models**. Panels **j, k**, and **l** present Kaplan–Meier survival curves stratified by median predicted 5-year risk from the reduced-50% models on the held-out test cohort for cirrhosis, all-cause mortality, and HCC, respectively, with log-rank tests indicating significant separation of risk groups. Participant counts less than 20 are censored in accordance with the All of Us Research Program Data and Statistics Dissemination Policy to protect participant privacy.

In addition to continuous time-to-event prediction, we binarily stratified the patients at 3-, 5-, and 10-year horizons using the top 50% reduced models. Test-set AUROC values for cirrhosis were 0.6953, 0.6815, and 0.6822 at 3, 5, and 10 years, respectively. For HCC, AUROC values were 0.6771, 0.7322, and 0.7388, and for all-cause mortality, 0.7812, 0.8053, and 0.7858. Time-dependent ROC curves are presented in Fig. 2(**g, h, i**). AUROC results for all additional experiments are provided in Supplementary Tables 14, 16, and 18.

Risk stratification based on median predicted 5-year risk from the reduced models yielded distinct survival separation for each outcome. Kaplan–Meier curves demonstrated significant differences between high- and low-risk groups (log-rank p < 0.01) for cirrhosis, HCC, and mortality (Fig. 2(**j, k, l**)). Comparable stratification patterns were observed for full-feature and top 25% reduced models (Supplementary Fig. 3). Calibration metrics for all feature sets are summarized in Supplementary Table 20.

### Contribution and behavior of baseline features across outcomes

The reduced models retained 21 features selected through cross-validated permutation importance and reevaluated using SHAP on the held-out test sets. These included age at CHC diagnosis, deprivation index, liver enzymes (ALT, AST, ALP), markers of hepatic function (albumin, total bilirubin), metabolic indicators (A1c, triglycerides, LDL), blood pressure measures, major comorbidities, selected medication exposures, and several chromosome 19 and 22 variants. The recurrence of these features across outcomes indicates shared clinical pathways linking chronic hepatitis C to cirrhosis, HCC, and mortality.

Age at CHC diagnosis was the dominant predictor across all outcomes (Fig. 3(**a, d, i**)), with the largest absolute SHAP magnitudes. Dependency analyses showed a nonlinear gradient, with risk increasing more sharply beyond approximately 50– 55 years, particularly for HCC and mortality (Fig. 3(**b, c**)). Elevated ALT (Fig. 3**g**) and AST (Fig. 3**h**) demonstrated monotonic positive associations with cirrhosis and HCC, while lower albumin (Fig. 3**l**) and higher bilirubin (Fig. 3**m**) were more strongly associated with mortality. Metabolic features and medication exposures contributed modest but consistent effects, especially in mortality models, reflecting systemic disease burden.

**Fig. 3.**
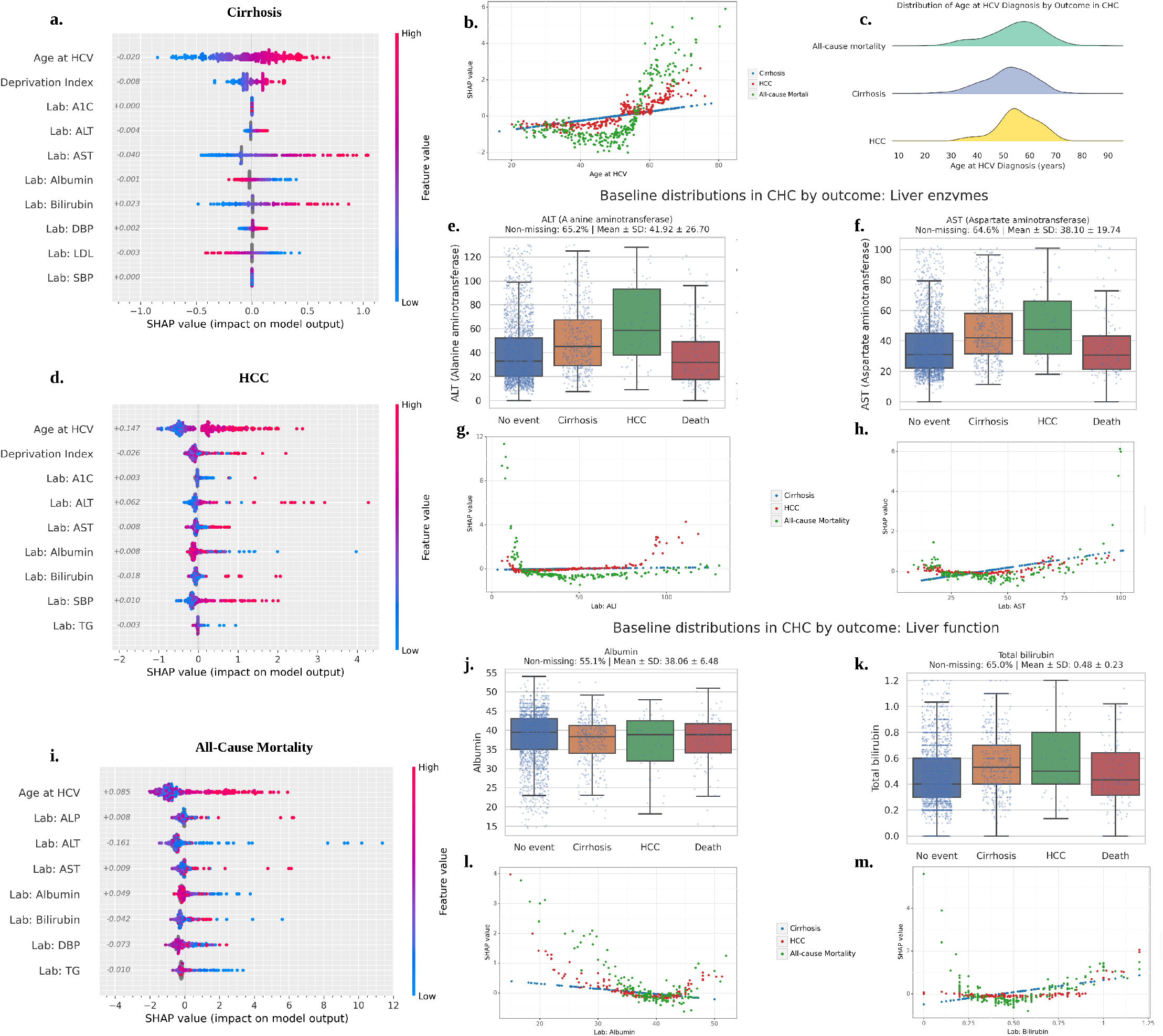
Contribution and behavior of numeric baseline features across outcomes. This figure summarizes how numeric baseline variables influence model predictions and outcomes in chronic hepatitis C (CHC), using held-out test data from the best-performing model for each endpoint (Cirrhosis: Coxnet-LASSO; Hepatocellular carcinoma (HCC) and all-cause mortality: Random Survival Forest). **Global importance of numeric features**. Panels **a, d**, and **i** show SHAP summary plots for cirrhosis, HCC, and all-cause mortality, respectively, highlighting the direction and magnitude of impact for key numeric features, including age at HCV diagnosis, liver enzymes, albumin, bilirubin, and metabolic markers. Each point represents an individual patient, colored by feature value, illustrating heterogeneous contributions to predicted risk. **Age-related effects across outcomes**. Panel **b** presents SHAP dependency plots for age at HCV diagnosis stratified by outcome, demonstrating non-linear and outcome-specific risk trajectories, while panel **c** shows the corresponding baseline age distributions across outcome groups. **Liver enzyme dynamics and missingness patterns**. Panels **e** and **f** depict baseline distributions of *alanine aminotransferase* (ALT) and *aspartate aminotransferase* (AST), respectively, across outcome groups, including proportions of non-missing measurements. Panels **g** and **h** show SHAP dependency plots for ALT and AST, illustrating how both observed values and missingness patterns contribute to risk stratification. **Liver function markers**. Panels **j** and **k** summarize baseline distributions of serum albumin and total bilirubin across outcome groups, while panels **l** and **m** present their corresponding SHAP dependency plots.

Laboratory and vital measurements were summarized as mean values within ±180 days of CHC diagnosis. Missingness was substantially present for several metabolic markers, including A1c, triglycerides, HDL, and LDL (65–75%), whereas liver enzymes and albumin showed moderate missingness (approximately 35–45%) (Fig. 3**e, f, j, k**). In addition, on average, fewer observations were available to compute the mean of A1c, triglycerides, HDL, and LDL compared to the other labs and vitals (Supplementary Table 9). Among the evaluated strategies for handling missing values, median imputation combined with explicit missing indicators consistently yielded better or comparable cross-validated C-index performance and was therefore adopted for subsequent analysis.

Genomic variants demonstrated outcome-specific and genotype-dependent effects (Supplementary Fig. 5a–c). In cirrhosis, *chr*19 : 19268740 showed opposing contributions by genotype, with homozygous alternate carriers exhibiting a strong negative mean SHAP (approximately −0.4). In contrast, HCC and mortality models showed substantially larger magnitudes. Homozygous alternate genotypes at *chr*22 : 43928850 and *chr*22 : 43928847 in HCC exceeded +1.0 mean SHAP, and *chr*19 : 19268740 and *chr*19 : 39252525 in mortality displayed stepwise increases approaching or exceeding +1.0. These results indicate that selected chromosome 19 and 22 loci exert meaningful and graded effects, particularly for HCC and mortality risk.

Medication exposures were among the strongest non-laboratory predictors (Supplementary Fig. 5d–f). In cirrhosis, diabetes and related therapies contributed positively (approximately +0.3 to +0.4), while metformin showed a negative association. In HCC, insulin use demonstrated a moderate positive contribution (approximately +0.4). The largest effects were observed in mortality models, where insulin use exceeded +3.0 mean SHAP and antihypertensive therapy approached +2.0, substantially larger than in liver-specific models. These findings suggest that treatment intensity and cardiometabolic burden are particularly informative for mortality risk stratification.

Patient-level SHAP waterfall analyses demonstrated biologically coherent risk attribution across outcomes. True positive cases were driven by strong positive contributions from elevated AST, ALT, bilirubin, older age at CHC diagnosis, and cardiometabolic comorbidities (Supplementary Fig. 6a, e, i). True negative cases showed dominant protective effects, including younger age and preserved liver synthetic function (Supplementary Fig. 6b, f, j). False negatives were characterized by largely protective laboratory and clinical profiles despite eventual events, indicating limited measurable risk within the feature window (Supplementary Fig. 6d, h, l). False positives, particularly for HCC and mortality, exhibited substantial age, laboratory, or medication-related risk signals in individuals who were censored or event-free, reflecting inherent uncertainty in time-to-event prediction (Supplementary Fig. 6c, g, k). Overall, individual explanations aligned with known disease mechanisms and reinforced model interpretability.

### Demographic heterogeneity and fairness of model performance

The cohort demonstrated limited socioeconomic separation across outcomes. The mean deprivation index was 0.33 ± 0.06 (median 0.31; IQR 0.29 − 0.37), with nearly identical distributions among cirrhosis and HCC cases and slightly higher values among those who died. Descriptive gradients of event proportions across DI strata showed modest, non-monotonic variation across race and ethnicity (Fig. 4(**a, d, i, l**)), without clear population-level stratification by deprivation alone.

**Fig. 4.**
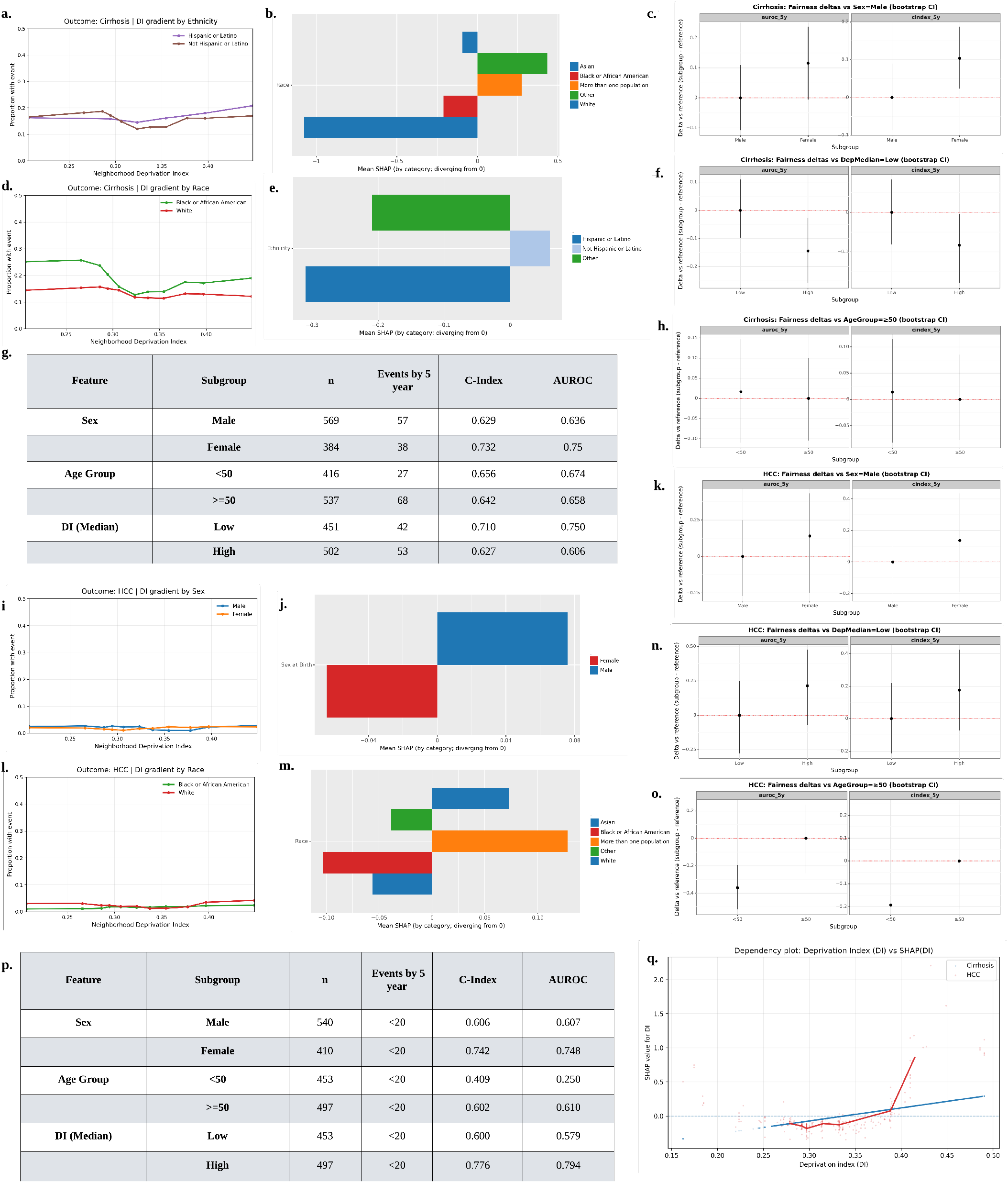
Demographic patterns, subgroup performance, and fairness across cirrhosis and HCC outcomes. This figure integrates descriptive epidemiology, model-based interpretation, subgroup discrimination, and fairness analyses for cirrhosis (Coxnet–LASSO) and hepatocellular carcinoma (HCC; Random Survival Forest), evaluated on the held-out test dataset. For cirrhosis (panels **a–h**): deprivation index (DI) gradients by ethnicity and race (**a**,**d**), category-level SHAP contributions for race and ethnicity (**b**,**e**), subgroup-specific discrimination metrics (C-index and AUROC at 5 years) across sex, age group, and median DI strata (**g**), and bootstrap-based fairness deltas for AUROC_5*y*_ and C-index_5*y*_ comparing sex, DI (median split), and age groups (**c**,**f**,**h**). For HCC (panels **i–o**): analogous DI gradients (**i**,**l**), SHAP contributions for sex and race (**j**,**m**), subgroup-specific performance metrics (**p**), and bootstrap fairness deltas for sex, DI, and age groups (**k**,**n**,**o**). Panel **q** presents a joint dependency plot of SHAP(DI) versus DI for cirrhosis and HCC, illustrating the direction and magnitude of DI-associated risk contributions across outcomes. Across models, discrimination remains stable with limited evidence of systematic performance disparities across demographic subgroups. Participant counts less than 20 are censored in accordance with the All of Us Research Program Data and Statistics Dissemination Policy to protect participant privacy.

Within predictive models, DI remained among the top reduced features for cirrhosis and HCC. SHAP dependency analysis showed a mild, approximately linear increase in cirrhosis risk with higher DI, whereas for HCC the association was weaker and more apparent at higher deprivation levels (Fig. 4**q**). Demographic variables, including race and sex, demonstrated directional but smaller SHAP magnitudes relative to age and liver-related laboratory features. For cirrhosis and HCC, race was in the top 50% feature list; black and white races exhibited negative mean SHAP contribution. In HCC, male sex showed positive mean SHAP while in cirrhosis, non hispanic had positive mean SHAP.

Subgroup discrimination analyses in the held-out test sets showed stable performance across sex, age, and DI strata. For cirrhosis (Fig. 4**g**), females showed higher discrimination than males (C-index 0.732 vs 0.629; AUROC 0.750 vs 0.636), and low-DI groups had slightly higher AUROC than high-DI groups (0.750 vs 0.606), with comparable performance across age groups. For HCC (Fig. 4**p**)), females again demonstrated higher discrimination (C-index 0.742 vs 0.606; AUROC 0.748 vs 0.607), and the high-DI group showed numerically higher performance than the low-DI group. However, bootstrap fairness deltas for C-index5*y* and AUROC5*y* were centered around zero with confidence intervals spanning zero across sex, age, and DI strata (Fig. 4(**c, f, h, k, n, o**)), indicating no statistically significant performance disparities.

### Disease trajectories and competing risks

To further validate model-derived feature attributions, we compared signed mean SHAP values with Cox proportional hazards coefficients for each outcome (Fig. 5(**a, b**)). Across cirrhosis, HCC, and all-cause mortality, there was clear directional concordance between SHAP contributions and log-hazard estimates. Age at CHC diagnosis demonstrated consistent positive associations in both frameworks, as did key liver-related markers. For example, elevated ALT and AST showed positive Cox coefficients that aligned with positive SHAP contributions in cirrhosis and HCC models, whereas lower albumin and higher bilirubin were more strongly associated with mortality risk.

**Fig. 5.**
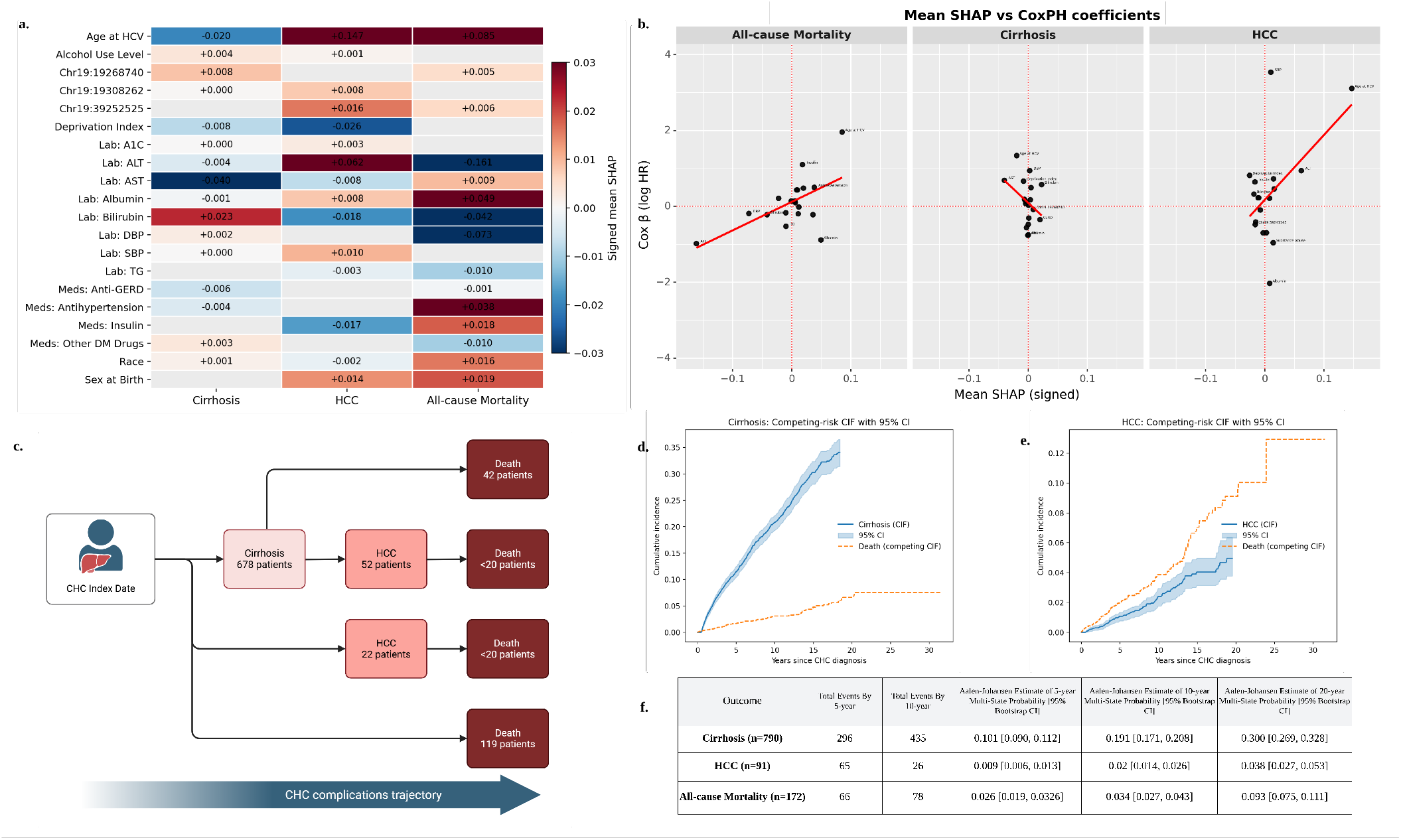
Integrated feature importance, disease progression, and competing-risk outcomes in chronic hepatitis C (CHC). **Panel a**. Heatmap of signed mean SHAP values for the reduced top-50% feature sets across cirrhosis, hepatocellular carcinoma (HCC), and all-cause mortality, summarizing consistent and outcome-specific predictors identified by the best-performing models (Coxnet-LASSO for cirrhosis; Random Survival Forest for HCC and all-cause mortality) on the held-out test dataset. **Panel b**. Scatter plots comparing signed mean SHAP values (x-axis) with Cox proportional hazards coefficients (y-axis), demonstrating concordance between model-agnostic feature importance and traditional log-hazard estimates across outcomes. **Panel c**. Schematic representation of observed patient trajectories from the CHC index date, illustrating progression pathways to cirrhosis, HCC, and death and their relative frequencies in the cohort. **Panels d–e**. Cumulative incidence functions (CIFs) with 95% confidence intervals for cirrhosis (**d**) and HCC (**e**), accounting for death as a competing event. **Panel f**. Multi-state Aalen–Johansen estimates of cumulative incidence at 5, 10, and 20 years for cirrhosis, HCC, and all-cause mortality. Participant counts less than 20 are censored in accordance with the All of Us Research Program Data and Statistics Dissemination Policy to protect participant privacy.

We then evaluated longitudinal outcome dynamics using competing-risk analyses. Cumulative incidence functions (Fig. 5(**d, e**)) demonstrated substantially higher cumulative incidence for cirrhosis compared with HCC over follow-up, while death remained a persistent competing event for HCC. Fine– Gray subdistribution hazard estimates are provided in the Supplementary Fig. 6.

To contextualize these risks within disease progression, we examined observed patient trajectories and multi-state transitions (Fig. 5(**c, f**)). Progression from CHC to cirrhosis represented the most frequent pathway, with a smaller subset developing HCC either after cirrhosis or directly from CHC. Death occurred both subsequent to liver complications and independently. Multi-state Aalen–Johansen estimates quantified state occupancy over time, showing progressive accumulation of cirrhosis, comparatively lower but increasing HCC probability, and rising all-cause mortality with extended follow-up.

## Discussion

In this study, we developed and validated an interpretable survival modeling framework to predict progression to cirrhosis, hepatocellular carcinoma (HCC), and all-cause mortality in individuals with chronic hepatitis C (CHC) within a large, diverse national biobank. Using harmonized real-world data, we integrated survival modeling, feature reduction, time-horizon discrimination, subgroup robustness, interpretability analyses, and competing-risk trajectory modeling into a unified framework.

By integrating multimodal features, including demographic characteristics, comorbidities, medication exposures, laboratory biomarkers, socioeconomic context, and selected germline variants, our models achieved consistent discrimination across internal validation and held-out testing. Notably, penalized Cox regression performed competitively for cirrhosis, while ensemble tree-based methods demonstrated consistent performance for HCC and mortality. The neural Cox-nnet v2 model did not uniformly outperform classical or ensemble approaches, underscoring that increased architectural complexity does not guarantee improved survival discrimination in moderately sized clinical cohorts. Across outcomes, reduced models retaining only top 25% and top 50% of features retained discrimination comparable to full-feature models, demonstrating that most prognostic signal is concentrated within a compact set of routinely available baseline variables.

A central finding is that parsimonious models preserved predictive performance despite substantial feature compression. In the test set, reduction to the top 50% of features resulted in minimal change in discrimination (-0.22% for cirrhosis, +3.51% for HCC, and -0.81% for mortality). Even with the top 25% of features, performance remained stable, with absolute decreases below 2% for cirrhosis and mortality and a 0.95% increase for HCC. The reduced models comprised clinically interpretable domains, including age at CHC diagnosis, liver injury markers (ALT, AST), hepatic reserve indicators (albumin, bilirubin), cardiometabolic comorbidities and related medications, blood pressure measures, deprivation index, and selected chromosome 19 and 22 variants. These findings support the feasibility of implementing risk stratification using a focused, clinically meaningful feature set rather than high-dimensional black-box representations.

Model explanations reinforced existing known clinical and biological risk factors for liver complications. Age consistently anchored baseline vulnerability, with nonlinear escalation at older ages ^10–12^. Markers of hepatocellular injury were more strongly associated with cirrhosis and HCC ^41–43^, whereas markers of hepatic synthetic function and systemic cardiometabolic burden were more influential for mortality ^44–46^. This divergence highlights that liver-specific progression and overall survival reflect overlapping but distinct biological processes ^47–49^. Genotype-dependent, graded contributions at selected loci were observed, with stepwise shifts in predicted risk across allele dosages (0/1/2). Importantly, this pattern reflects the statistical encoding of genotype dosage rather than an a priori assumption of additive biological effects. In our framework, variants were modeled using reference (REF) and alternative (ALT) allele dosage without pre-specifying effect direction. The designation of an allele as “reference” or “alternative” reflects alignment to a reference genome build rather than pathogenicity, population frequency, or ancestral status. It is therefore entirely possible for the ALT allele to represent the ancestral or more common population allele, while the REF allele may be rarer or deleterious in specific contexts. Likewise, homozygosity for the ALT allele (dosage = 2) does not inherently imply increased biological burden; it may correspond to the healthy or population-predominant state. The observed graded SHAP contributions thus indicate learned monotonic associations within this cohort rather than confirmation of strictly additive genetic architecture. The larger magnitudes observed for HCC and mortality compared with cirrhosis suggest that host genetic background may contribute more strongly to carcinogenesis and systemic vulnerability than to fibrosis progression alone. Although exploratory and limited to a targeted variant set, these findings align with evidence that inherited susceptibility contributes to heterogeneity in clinical outcomes beyond stage-based proxies ^15,18–20,36^.

Assessing the internal validity of the model through patient-level SHAP analyses demonstrated coherent and clinically plausible risk attribution. True positive cases were driven by biologically consistent combinations of older age, elevated markers of hepatic injury, impaired synthetic function, and increased cardiometabolic burden, whereas true negatives were characterized predominantly by protective laboratory profiles and lower baseline vulnerability. False positive and false negative classifications reflected the inherent uncertainty of longitudinal risk prediction and competing-event dynamics rather than implausible feature contributions. Collectively, these findings support the mechanistic plausibility, stability, and interpretability of the modeling framework.

Importantly, antiviral treatment exposure was captured for only a minority of patients within the baseline feature window. Our findings should therefore be interpreted as baseline risk stratification rather than definitive post-sustained virologic response (SVR) residual risk estimation. However, this scenario mirrors real-world EHR constraints, where treatment may occur outside the observation window or be incompletely recorded ^33,34^. Within this context, our models demonstrate that baseline clinical phenotype still stratifies risk for cirrhosis, HCC, and mortality independent of stage categorization or fully captured SVR status. Even when fibrosis stage or treatment history is uncertain, routinely available clinical signals provide substantial prognostic information.

The competing-risk and multistate analyses contextualize these predictions within real-world CHC trajectories. Cirrhosis represented the most frequent transition, HCC accumulated more gradually ^50^, and death remained a persistent competing event, particularly for HCC ^26^. This has direct clinical implications. Staging-based guidelines define surveillance eligibility but do not quantify individualized risk magnitude nor account for competing mortality ^51^. Our results support a more nuanced approach in which surveillance intensity is informed not only by fibrosis proxies but also by patient-level risk phenotype and competing risk profile. Within the same fibrosis stage (F3/F4) category, one individual may demonstrate high long-horizon HCC risk with low competing mortality, which supports intensive surveillance. Another individual may have a substantial competing mortality risk ^52^, underscoring the importance of patient-centered, goal-directed care ^53^.

Demographic variables such as sex and race contributed modest directional effects without driving discrimination. Performance remained stable across sex, age, and deprivation strata, with no statistically significant subgroup disparities in discrimination metrics. While demographic features may reflect contextual and exposure-related risk ^54,55^, the predictive framework did not exhibit measurable performance imbalance within this cohort. Ongoing external validation and calibration monitoring remain essential before clinical deployment.

This study has several notable strengths. The use of the All of Us (AoU) Research Program enabled evaluation within a geographically and demographically diverse, nationally-representative cohort with harmonized clinical, laboratory, social, medication, and genomic data, enhancing representativeness beyond traditional single-center liver cohorts. We framed disease progression explicitly as a time-to-event process, incorporating right censoring and competing risks to align analysis with the longitudinal nature of chronic liver disease. A transparent and reproducible modeling pipeline compared penalized Cox models, ensemble machine learning methods, and neural survival approaches within a consistent cross-validation and held-out testing framework. Importantly, substantial feature reduction preserved discrimination (≤ 2 % absolute change in test C-index for cirrhosis and mortality and improvement for HCC), demonstrating that prognostic signal is concentrated within a compact, clinically interpretable feature set. The integration of permutation-based selection and SHAP interpretability strengthened biological plausibility, while subgroup fairness analyses and robustness checks supported stability across demographic and socioeconomic strata. Concordance between SHAP attributions and Cox coefficients further indicated alignment with established pathophysiology rather than opaque algorithmic artifacts.

Several limitations merit consideration. The retrospective observational design introduces potential misclassification and residual confounding inherent to EHR-derived phenotypes. Antiviral treatment exposure was incompletely captured within the baseline window, and findings should therefore be interpreted as baseline risk stratification rather than definitive post-SVR residual risk estimation. The ±180-day feature window may not fully capture cumulative or evolving disease burden. Missingness was substantial for certain metabolic markers, and although missingness indicators improved performance, informative missingness cannot be excluded. Genomic analyses were limited to a targeted set of variants and require replication. Event counts for HCC were modest, potentially constraining model stability and partially explaining comparable performance of simpler and ensemble approaches. Finally, external validation beyond the AoU infrastructure and prospective evaluation of clinical utility are necessary before translation into routine care. Future work should also explore dynamic updating of risk over time, temporal shifts in clinical management, integration of longitudinal laboratory trajectories, and assessment of clinical utility in decision-analytic frameworks.

In summary, we developed and internally validated an interpretable, multimodal survival modeling framework that quantifies heterogeneous risk of cirrhosis, HCC, and mortality in chronic hepatitis C using routinely available baseline data. Reduced-feature models preserved discrimination while enhancing interpretability, and competing-risk and multistate analyses contextualized predictions within realistic disease trajectories. Rather than replacing fibrosis staging, this approach refines it by quantifying individualized risk magnitude and incorporating competing mortality considerations. With further validation, such parsimonious and biologically coherent risk stratification may support more personalized surveillance intensity, comorbidity optimization, and longitudinal care planning for individuals living with chronic hepatitis C.

## Data Availability

This study used data from the All of Us Research Program's Control Tier Dataset (Controlled Tier) v8, available to authorized users on the Researcher Workbench.

## Code availability

All analyses were conducted within the All of Us Researcher Workbench (Controlled Tier v8) using the platform’s managed Python environment (version 3.9). Cohort extraction was performed using SQL, and data preprocessing and modeling were implemented in Python using `pandas, NumPy, scikit-learn,` and `scikit-survival`. Neural network–based survival models were developed with PyTorch. Codes used for the analysis are available at https://github.com/hikf3/survpipe.

## Acknowledgements

We thank our colleagues at the Institute for Population and Precision Health (IPPH), University of Chicago, for their support and insightful discussions throughout this project. This research was conducted using data from the All of Us Research Program, and we gratefully acknowledge the participants who contributed their data to advance biomedical research.

## Author contributions

H.I. conceived the study, performed the data analysis, developed the computational pipeline, and drafted the manuscript. A.A. contributed to data processing, model validation, interpretation of results, and manuscript preparation. J.F. contributed conceptual guidance and reviewed and validated the study design, results, and interpretations. H.A. supervised the project, provided conceptual guidance and critical revisions, and oversaw the overall study design. All authors reviewed, revised, and approved the final manuscript.

## Additional information

The supplementary document contains additional tables and graphical illustrations to support the analysis of the manuscript.

## Supplementary Document

### 1 Cohort and Outcome Definition

- **Define the time origin (CHC index date)**. For each participant, we used the first recorded chronic hepatitis C diagnosis date (`first_hepc_datetime`) as time zero for all time-to-event calculations. All subsequent event and censoring times were measured relative to this index date.
- **Construct the administrative follow-up end date from EHR conditions**. Using the CHC cohort’s condition records (`hepc_conditions`), we computed a participant-level last-observed clinical date (`followup_ end_from_conditions`) as the maximum of `condition_end_datetime` when available, other-wise `condition_start_datetime`. Participants with follow-up dates occurring on or before the CHC index date were set to missing follow-up.
- **Derive event indicators and event timestamps for liver outcomes**. For each outcome (cirrhosis, liver cancer, and death), we used cohort-level flags (`has_{outcome}`) as binary event indicators and the corresponding first event date (`first_{outcome}_datetime`) as the event timestamp.
- **Compute time-to-event with right censoring**. For each outcome, we calculated:
  - event time in days: *t*_event_ = `first_{outcome}_datetime − first_hepc_datetime,`
  - censoring time in days: *t*_cens_ = `followup_end_time − first_hepc_datetime,`
  - observed time: *t* = *t*_event_ if `has_{outcome}` = 1, otherwise *t* = *t*_cens_. Times were stored in both days (`time_{outcome}_days`) and years (`time_{outcome}_years` = *t/*365.25). For instances where `first_{outcome}_datetime < first_hepc_datetime`, resulting in negative durations, were excluded from the cohort.
- **Ascertain all-cause mortality and time-to-death**. We extracted death dates from the AoU death table (`aou_death`) and retained the earliest recorded death per participant. A death indicator was defined as `event_death` = 1 if a death date was present. Time-to-death was computed from the CHC index date using an end date defined as death date for decedents and `followup_end_from_conditions` for non-decedents, yielding `days_to_death` and `years_to_death`. Invalid times (missing or negative) were set to missing and death was treated as not observed (censored).
- **Competing-risk adjustment for liver outcomes (death-aware censoring)**. To correctly handle death occurring prior to a liver outcome, we constructed death-aware follow-up for each outcome by defining the observed time as the minimum of three candidate times: the outcome time, the death time, and the administrative censoring time. We then defined an adjusted event indicator that counted the outcome only if the outcome time equaled this minimum time (i.e., occurred before death and before administrative censoring). These were stored as `time_{outcome}_min_years` and `event_{outcome}_adj`, while retaining the original outcome time and indicator columns unchanged.

**Supplementary Table 1:**
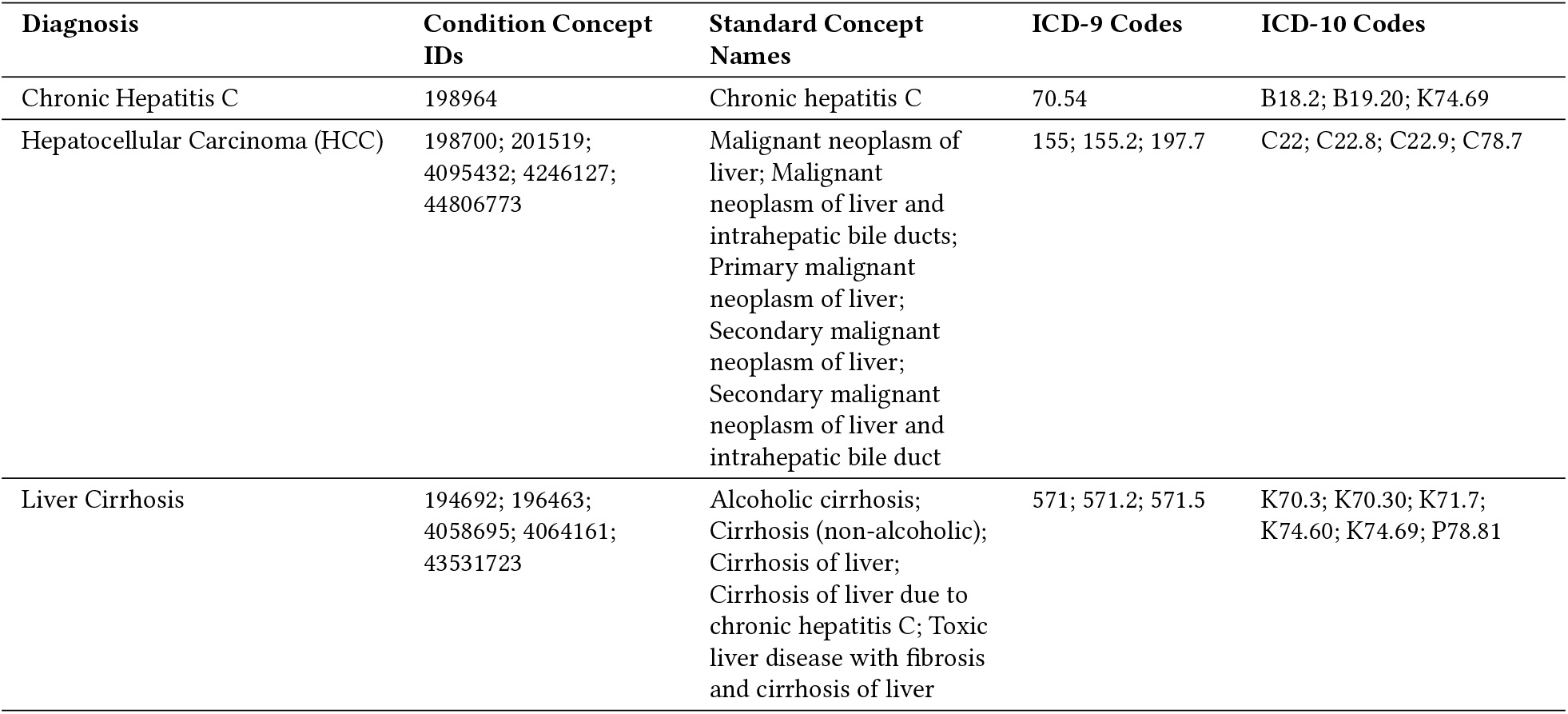
Cohort and outcome definitions with corresponding ICD codes used in the study cohort.

### 2. Feature extraction in *All of Us*

#### 2.1 Comorbidities

**Supplementary Table 2:**
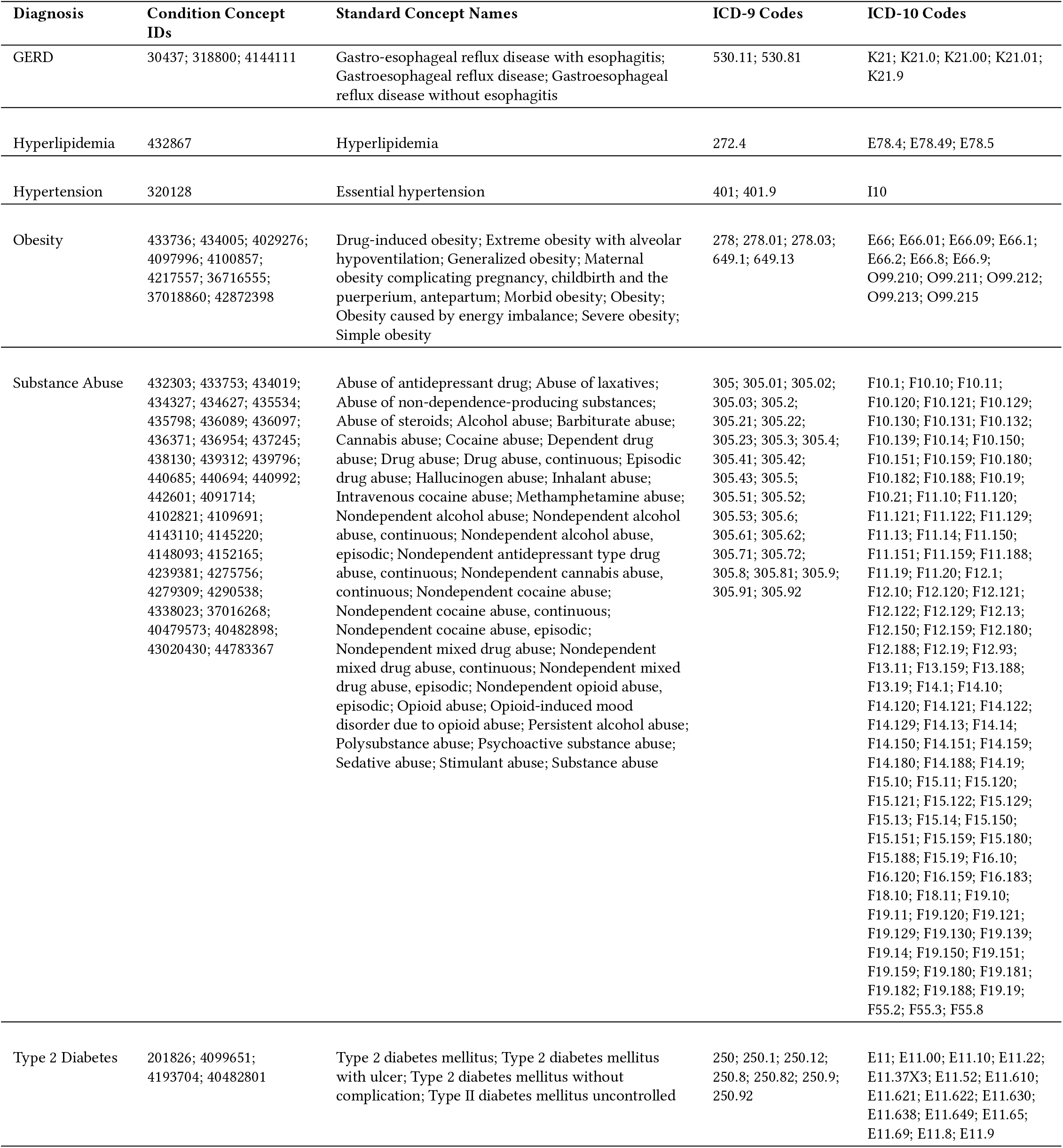
Comorbidity definitions and ICD mappings used for condition-based feature extraction in All of Us.

| Diagnosis | Condition Concept IDs | Standard Concept Names | ICD-9 Codes | ICD-10 Codes |
| --- | --- | --- | --- | --- |
| GERD | 30437; 318800; 4144111 | Gastro-esophageal reflux disease with esophagitis; Gastroesophageal reflux disease; Gastroesophageal reflux disease without esophagitis | 530.11; 530.81 | K21; K21.0; K21.00; K21.01; K21.9 |
| Hyperlipidemia | 432867 | Hyperlipidemia | 272.4 | E78.4; E78.49; E78.5 |
| Hypertension | 320128 | Essential hypertension | 401; 401.9 | I10 |
| Obesity | 433736; 434005; 4029276; 4097996; 4100857; 4217557; 36716555; 37018860; 42872398 | Drug-induced obesity; Extreme obesity with alveolar hypoventilation; Generalized obesity; Maternal obesity complicating pregnancy, childbirth and the puerperium, antepartum; Morbid obesity; Obesity; Obesity caused by energy imbalance; Severe obesity; Simple obesity | 278; 278.01; 278.03; 649.1; 649.13 | E66; E66.01; E66.09; E66.1; E66.2; E66.8; E66.9; O99.210; O99.211; O99.212; O99.213; O99.215 |
| Substance Abuse | 432303; 433753; 434019; 434327; 434627; 435534; 435798; 436089; 436097; 436371; 436954; 437245; 438130; 439312; 439796; 440685; 440694; 440992; 442601; 4091714; 4102821; 4109691; 4143110; 4145220; 4148093; 4152165; 4239381; 4275756; 4279309; 4290538; 4338023; 37016268; 40479573; 40482898; 43020430; 44783367 | Abuse of antidepressant drug; Abuse of laxatives; Abuse of non-dependence-producing substances; Abuse of steroids; Alcohol abuse; Barbiturate abuse; Cannabis abuse; Cocaine abuse; Dependent drug abuse; Drug abuse; Drug abuse, continuous; Episodic drug abuse; Hallucinogen abuse; Inhalant abuse; Intravenous cocaine abuse; Methamphetamine abuse; Nondependent alcohol abuse; Nondependent alcohol abuse, continuous; Nondependent alcohol abuse, episodic; Nondependent antidepressant type drug abuse, continuous; Nondependent cannabis abuse, continuous; Nondependent cocaine abuse; Nondependent cocaine abuse, continuous; Nondependent cocaine abuse, episodic; Nondependent mixed drug abuse; Nondependent mixed drug abuse, continuous; Nondependent mixed drug abuse, episodic; Nondependent opioid abuse, episodic; Opioid abuse; Opioid-induced mood disorder due to opioid abuse; Persistent alcohol abuse; Polysubstance abuse; Psychoactive substance abuse; Sedative abuse; Stimulant abuse; Substance abuse | 305; 305.01; 305.02; 305.03; 305.2; 305.21; 305.22; 305.23; 305.3; 305.4; 305.41; 305.42; 305.43; 305.5; 305.51; 305.52; 305.53; 305.6; 305.61; 305.62; 305.71; 305.72; 305.8; 305.81; 305.9; 305.91; 305.92 | F10.1; F10.10; F10.11; F10.120; F10.121; F10.129; F10.130; F10.131; F10.132; F10.139; F10.14; F10.150; F10.151; F10.159; F10.180; F10.182; F10.188; F10.19; F10.21; F11.10; F11.120; F11.121; F11.122; F11.129; F11.13; F11.14; F11.150; F11.151; F11.159; F11.188; F11.19; F11.20; F12.1; F12.10; F12.120; F12.121; F12.122; F12.129; F12.13; F12.150; F12.159; F12.180; F12.188; F12.19; F12.93; F13.11; F13.159; F13.188; F13.19; F14.1; F14.10; F14.120; F14.121; F14.122; F14.129; F14.13; F14.14; F14.150; F14.151; F14.159; F14.180; F14.188; F14.19; F15.10; F15.11; F15.120; F15.121; F15.122; F15.129; F15.13; F15.14; F15.150; F15.151; F15.159; F15.180; F15.188; F15.19; F16.10; F16.120; F16.159; F16.183; F18.10; F18.11; F19.10; F19.11; F19.120; F19.121; F19.129; F19.130; F19.139; F19.14; F19.150; F19.151; F19.159; F19.180; F19.181; F19.182; F19.188; F19.19; F55.2; F55.3; F55.8 |
| Type 2 Diabetes | 201826; 4099651; 4193704; 40482801 | Type 2 diabetes mellitus; Type 2 diabetes mellitus with ulcer; Type 2 diabetes mellitus without complication; Type II diabetes mellitus uncontrolled | 250; 250.1; 250.12; 250.8; 250.82; 250.9; 250.92 | E11; E11.00; E11.10; E11.22; E11.37X3; E11.52; E11.610; E11.621; E11.622; E11.630; E11.638; E11.649; E11.65; E11.69; E11.8; E11.9 |

#### 2.2 Medications

- **Concept-set definition**. For each medication group (e.g., Antibiotics, Anti-GERD), we defined a concept set as the union of the listed RxNorm concept identifiers (optionally including descendant concepts when concept hierarchy expansion was enabled).
- **Patient-level exposure flag (within** ±**180 days)**. For each patient, we set the binary indicator to 1 if the patient had ≥ 1 drug exposure record (e.g., in `drug_exposure`) whose `drug_concept_id` matched any concept in the corresponding concept set within a ±180-day window centered on the CHC diagnosis (index) date. Otherwise, the indicator was set to 0.
- **Multiple-drug collapse**. If a patient had exposures to multiple drugs within the same medication group (or repeated exposures to the same drug), these events were collapsed into a single presence/absence indicator to reduce feature sparsity and improve interpretability.
- **Non-mutually exclusive groups**. Medication indicators were not constrained to be mutually exclusive; a patient could simultaneously satisfy multiple exposures (e.g., `Antibiotics=1, Anti-GERD=1, and Insulin=1`), reflecting real-world polypharmacy.

**Supplementary Table 3:**
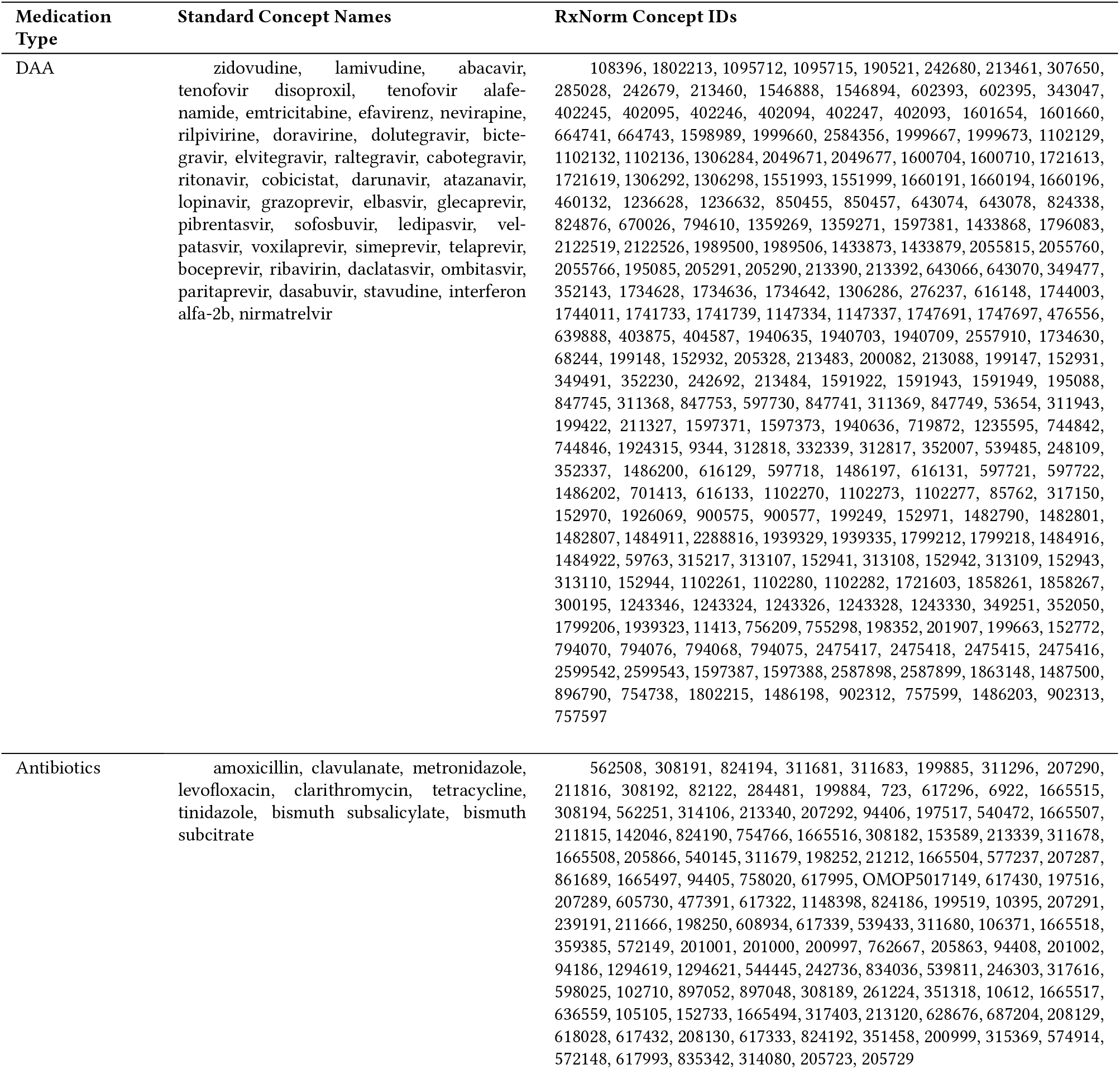
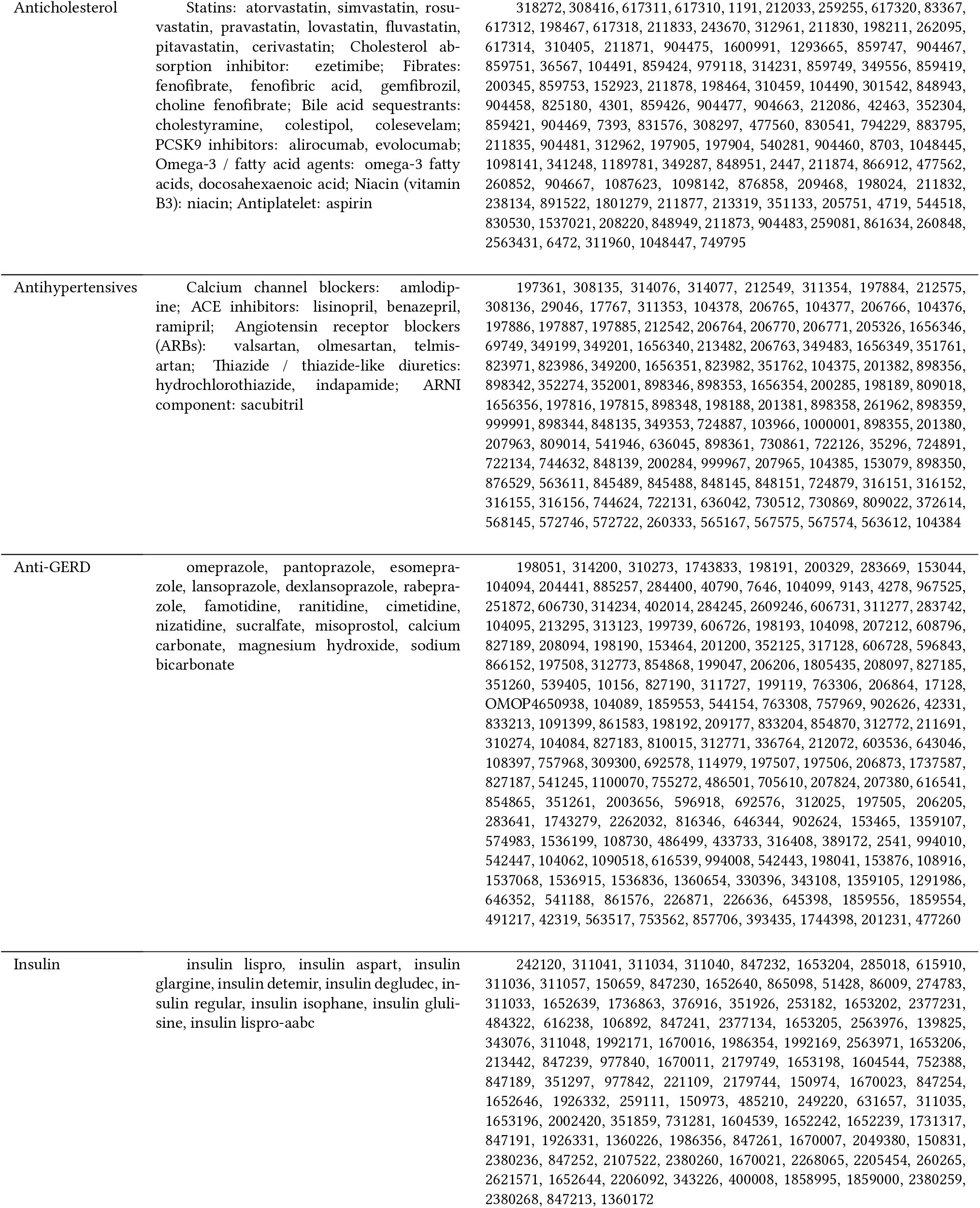
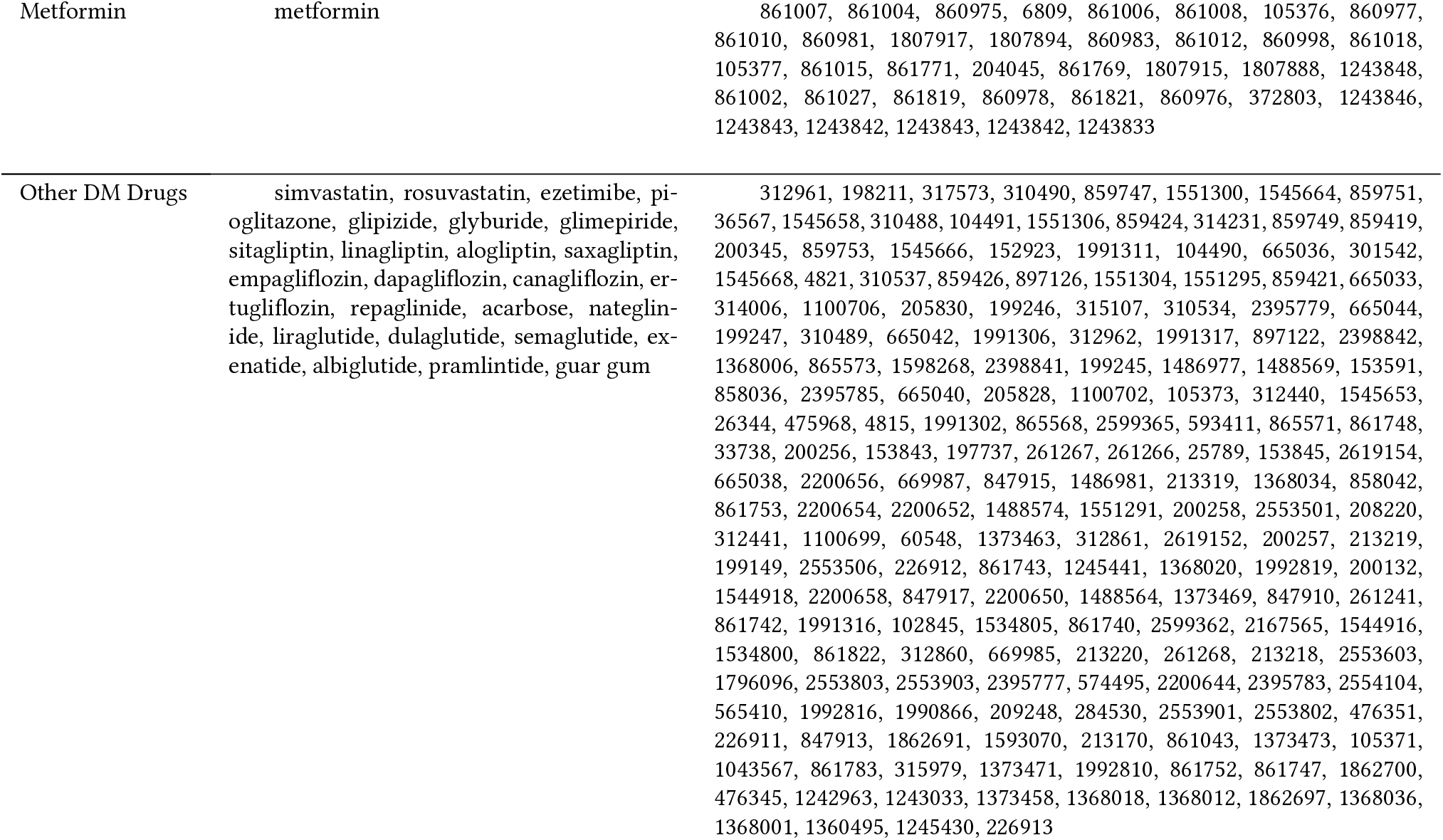
Medication group definitions used to construct binary exposure indicators. Each group aggregates RxNorm concept identifiers that map to therapeutically related medications.

#### 2.3 Lab measurements and vitals

- **Concept-set definition (laboratory measurements and vitals)**. For each laboratory analyte and vital sign (HDL, LDL, triglycerides, albumin, total bilirubin, ALT, AST, ALP, systolic blood pressure, and diastolic blood pressure), we defined a concept set based on OMOP `measurement_concept_ids` mapped to standard LOINC codes. All qualifying measurement records were extracted from the `measurement` table.
- **Temporal alignment and observation window**. Measurements were aligned to each participant’s chronic hepatitis C (CHC) diagnosis date (index date). Only observations occurring within a ±180-day window centered on the index date (− 180 < days < +180) were retained to ensure temporal relevance to baseline disease status.
- **Participant-level mean computation**. Participant-level summary values were computed using a robust, multi-step approach. If a single valid measurement was available within the window, that value was used directly. If multiple measurements were available, within-participant outliers were identified and removed using Tukey’s rule (1.5 × IQR), with the lower bound truncated at zero for non-negative analytes. The mean of the remaining measurements was then calculated.
- **Global outlier handling**. After participant-level means were derived, a second outlier screen was applied across the cohort using Tukey’s rule on the distribution of participant-level means. Extreme values were set to missing to limit undue influence of biologically implausible measurements.
- **Feature usage**. The resulting laboratory and vital sign summaries were retained as continuous features for downstream survival and predictive modeling analyses.

**Supplementary Table 4:** Laboratory and vital sign concept definitions and summary of observations used to compute participant-level means.

| Lab/Vital | Measurement Concept ID | Standard Concept Name | LOINC | Mean # Obs. | IQR |
| --- | --- | --- | --- | --- | --- |
| HDL | 3007070 | Cholesterol in HDL [Mass/volume] in Serum or Plasma | 2085-9 | 1.409338993 | 1 |
| LDL | 3009966 | Cholesterol in LDL [Mass/volume] in Serum or Plasma by Direct assay | 18262-6 | 1.414588529 | 1 |
| TG | 3022192 | Triglyceride [Mass/volume] in Serum or Plasma | 2571-8 | 1.405882353 | 1 |
| Albumin | 3024561 | Albumin [Mass/volume] in Serum or Plasma | 1751-7 | 5.735188510 | 4 |
| Bilirubin | 3007359 | Bilirubin.indirect [Mass/volume] in Serum or Plasma | 1971-1 | 6.390610894 | 4 |
| ALT | 3006923 | Alanine aminotransferase [Enzymatic activity/volume] in Serum or Plasma | 1742-6 | 3.947321954 | 3 |
| AST | 3013721 | Aspartate aminotransferase [Enzymatic activity/volume] in Serum or Plasma | 1920-8 | 3.909494232 | 3 |
| ALP | 3013721 | Aspartate aminotransferase [Enzymatic activity/volume] in Serum or Plasma | 1920-8 | 3.909494232 | 3 |
| SBP | 3004249 | Systolic blood pressure | 8480-6 | 29.521788660 | 15 |
| DBP | 3012888 | Diastolic blood pressure | 8462-4 | 28.881766380 | 15 |

#### 2.4 Demographics and socioeconomic status

- **Cohort-linked demographics extraction**. For the chronic hepatitis C (CHC) cohort, we used the AoU `demographics` table for (`CHC`) cohort containing `person_id, sex_at_birth, race, ethnicity, and date_of_birth`. We retained only these fields and collapsed to a single record per participant by de-duplicating on `person_id`.
- **Age at CHC diagnosis**. We appended each participant’s CHC index date (`first_hepc_datetime`) to the demographics table. We computed `age_at_hcv_diagnosis` as the time difference between `first_hepc_datetime and date_of_birth`, converted from days to years using 365.25. Negative ages (arising from inconsistent dates) were treated as missing (`NaN`).
- **Cleaning and harmonization of categorical demographics**. To improve interpretability and handle non-informative responses, we recoded selected values in `race, ethnicity, and sex_at_birth` into discrete categories with missingness indicators for responses such as `PMI: Skip, Prefer not to answer`, and other non-standard categories. The cleaned variables were stored as `race_clean, ethnicity_clean, and sex_at_birth_clean`. These were merged back into the participant-level analytic table by `person_id` and written to the final modeling dataset.
- **Neighborhood deprivation index**. Socioeconomic context was captured by extracting the deprivation index from the All of Us socioeconomic table. The deprivation index was identified by column name matching (`deprivation_index`, coerced to numeric, deduplicated to one value per participant, and merged into the analytic dataset using `person_id`.

#### 2.5 Lifestyle surveys

- **Lifestyle concept aggregation**. Lifestyle factors, including alcohol use and smoking status, were derived from multiple All of Us survey question concept identifiers capturing frequency, quantity, and behavioral history. All relevant survey responses were first harmonized at the participant level.
- **Rule-based category construction**. Responses from related survey items were merged using predefined clinical rules to form interpretable categorical variables. For alcohol use, information on lifetime use, past-year drinking frequency, average daily consumption, and binge drinking was combined to classify participants as *Never, Moderate, Extreme*, or *Unknown*. For smoking, lifetime cigarette exposure and current use of cigarettes or alternative tobacco products were integrated to define *Smoker, Non-smoker*, or *Unknown* categories.
- **Conflict resolution and prioritization**. When conflicting or incomplete responses were present, priority rules were applied to assign the most clinically informative category (e.g., prioritizing evidence of heavy exposure over moderate or missing responses). Participants with insufficient information across all relevant items were classified as *Unknown*.
- **Model-ready encoding**. The final lifestyle variables were encoded as categorical features and, where required, transformed into indicator variables for inclusion in downstream analyses.

**Supplementary Table 5:** Derivation of lifestyle features from All of Us lifestyle survey data.

| Feature | Question Concept IDs | Key Survey Items | Operational Rules | Derived Categories |
| --- | --- | --- | --- | --- |
| Alcohol use level | 1586198, 1586207, 1586201, 1586213 | Alcohol Participant; Drink Frequency Past Year; Average Daily Drink Count; 6+ Drinks Occurrence; PMI responses | Never: Alcohol Participant = No OR Drink Frequency = Never OR 6+ Drinks = Never in last year. Extreme: Average daily drinks $\geq 7$ OR 6+ drinks weekly or daily. Moderate: Average daily drinks 1–6 OR drinking less than 4 times per week without binge drinking. Unknown: Only PMI/skip responses or insufficient information. Priority order: Never → Extreme → Moderate → Unknown. | Never; Moderate; Extreme; Unknown |
| Smoking status | 1585873, 1585860, 1585857, 1585867, 1585864, 1586174, 1586177, 1586162, 1586159, 1586185, 1586182, 1585870, 1586169, 1586166 | 100 Cigs Lifetime; Smoke Frequency; Cigar, hookah, and e-cigarette participation and frequency; PMI responses | Smoker: 100 Cigs Lifetime = Yes OR any current smoking behavior (cigarette, cigar, hookah, or e-cigarette) reported as Every Day or Some Days. Non-smoker: 100 Cigs Lifetime = No OR all current smoking reported as Not At All. Unknown: Only PMI/skip responses or conflicting answers. | Smoker; Non-smoker; Unknown |

**Supplementary Table 6:** Genetic variants included in the analysis and their functional annotation.

| rsID | Chromosome | Position | REF | ALT | Type | Gene | Reference (PMID) |
| --- | --- | --- | --- | --- | --- | --- | --- |
| rs28929474 | chr14 | 94378610 | C | T | exonic | SERPINA1 | 38632349; 39572673 |
| rs58542926 | chr19 | 19268740 | C | T | exonic | TM6SF2 | 37709864; 38632349 |
| rs739846 | chr19 | 19308262 | G | A | intronic | SUGP1 | 38632349 |
| rs12980275 | chr19 | 39241143 | A | G | intronic | IL28B | 20060832; 28050240 |
| rs12979860 | chr19 | 39248147 | C | T | intronic | IFNL4 | 37709864; 20060832 |
| rs8099917 | chr19 | 39252525 | T | G | intergenic | IL28B | 20060832 |
| rs429358 | chr19 | 44908684 | T | C | exonic | APOE | 37709864; 39572673 |
| rs483082 | chr19 | 44912921 | G | T | intergenic | APOC1 | 38632349; 39572673 |
| rs738409 | chr22 | 43928847 | C | G | exonic | PNPLA3 | 28050240; 38632349 |
| rs738408 | chr22 | 43928850 | C | T | exonic | PNPLA3 | 37709864; 38632349 |

#### 2.6 Genomic features

- **Literature-based SNP selection**. Ten single nucleotide polymorphisms (SNPs) associated with liver fibrosis, cirrhosis, hepatocellular carcinoma, or related metabolic liver phenotypes in chronic hepatisis C and non-viral liver disease were selected from prior peer-reviewed studies, including genome-wide association studies (GWAS).
- **Reference genome harmonization**. Genomic coordinates reported in GRCh37 were converted to GRCh38 using the UCSC Lift Genome Annotations (LiftOver) tool to ensure consistency with the *All of Us* short-read whole-genome-sequencing (WGS) data release.
- **Allele confirmation and gene mapping**. Reference SNP Identification numbers (rsIDs), reference alleles, and alternate alleles were confirmed using dbSNP. Gene-level annotations were assigned based on canonical gene mappings.
- **Functional classification**. Variant functional class (exonic, intronic, or intergenic) was determined using Ensembl Variant Effect Predictor (VEP), based on the most severe annotated consequence for the modeled reference and alternative allele. The final set of curated variants, including genomic coordinates, alleles, functional classification, and literature references is summarized in Table 6.
- **Genomic data source**. Short-read WGS data from the *All of Us* Research Program Controlled Tier (v8), aligned to GRCh38 reference genome.
- **Cohort-specific VCF extraction**. Variant call files (VCFs) were generated using the All of Us Researcher Workbench genomic extraction workflow and organized into sex-stratified outputs (male and female).
- **Hail import and MatrixTable construction**. Extracted `*.vcf.gz` shards were imported into Hail and written as separate sex-specific MatrixTables.
- **Exact variant matching**. MatrixTables were filtered to retain only the curated SNPs using exact matching on chromosome, position, reference allele, and alternate allele.
- **Allele dosage computation**. For each participant–variant pair, allele dosage was computed as the number of alternate alleles using `GT.n_alt_alleles()`, yielding values in *{*0, 1, 2*}*.
- **Feature reshaping**. Dosages were exported in long format and reshaped into participant-by-variant wide matrices (variant identifier: `chr:position`) for downstream integration with clinical and laboratory features.

**Supplementary Figure 1.**
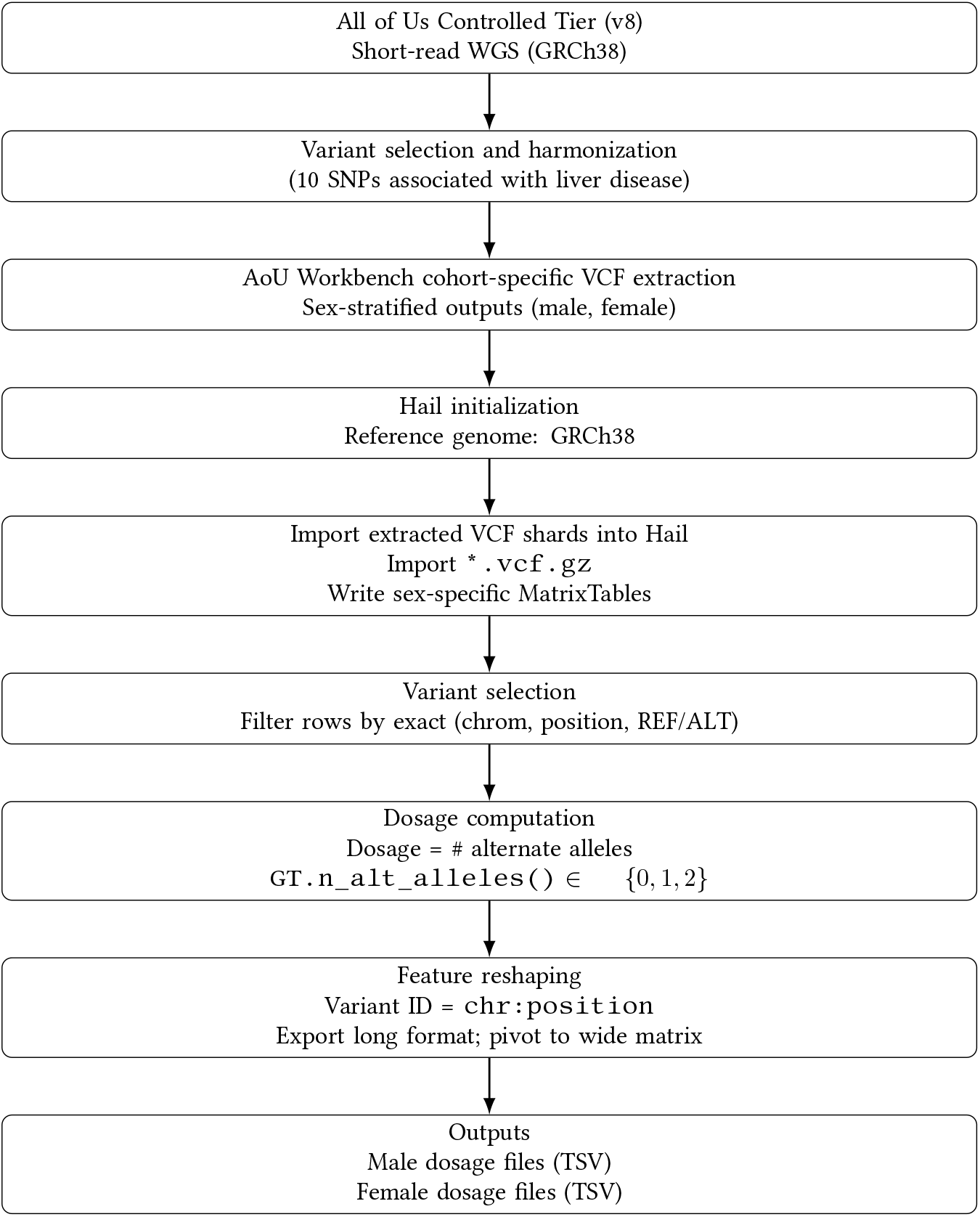
Genomic dosage derivation workflow.

### 3. Summary statistics

#### 3.1 Feature set

**Supplementary Table 7:** Feature sets (42 in full) used in the full survival prediction model.

| Outcomes | Categorical Feature Set | Numeric Feature Set |
| --- | --- | --- |
| • Cirrhosis<br>• Hepatocellular Carcinoma (HCC)<br>• All-cause Mortality | • Sex at birth<br>• Race<br>• Ethnicity<br>• Hypertension<br>• Type 2 diabetes<br>• Obesity<br>• Hyperlipidemia<br>• GERD<br>• Substance abuse<br>• Alcohol use level<br>• Smoking status<br>• Genetic variants<br>• DAA within 180 days<br>• Medication indicators: metformin, insulin, other diabetes drugs, antihypertensives, anticholesterol agents, antibiotics, anti-GERD | • Age at HCV diagnosis<br>• High-density lipoprotein (HDL), Low-density lipoprotein (LDL), triglycerides<br>• Hemoglobin A1c (HbA1c)<br>• Albumin, total bilirubin<br>• Alanine aminotransferase (ALT)<br>• Aspartate aminotransferase (AST)<br>• Alkaline phosphatase (ALP)<br>• Systolic and diastolic blood pressure<br>• Deprivation index |

#### 3.2 Population Characteristics

**Supplementary Table 8:**
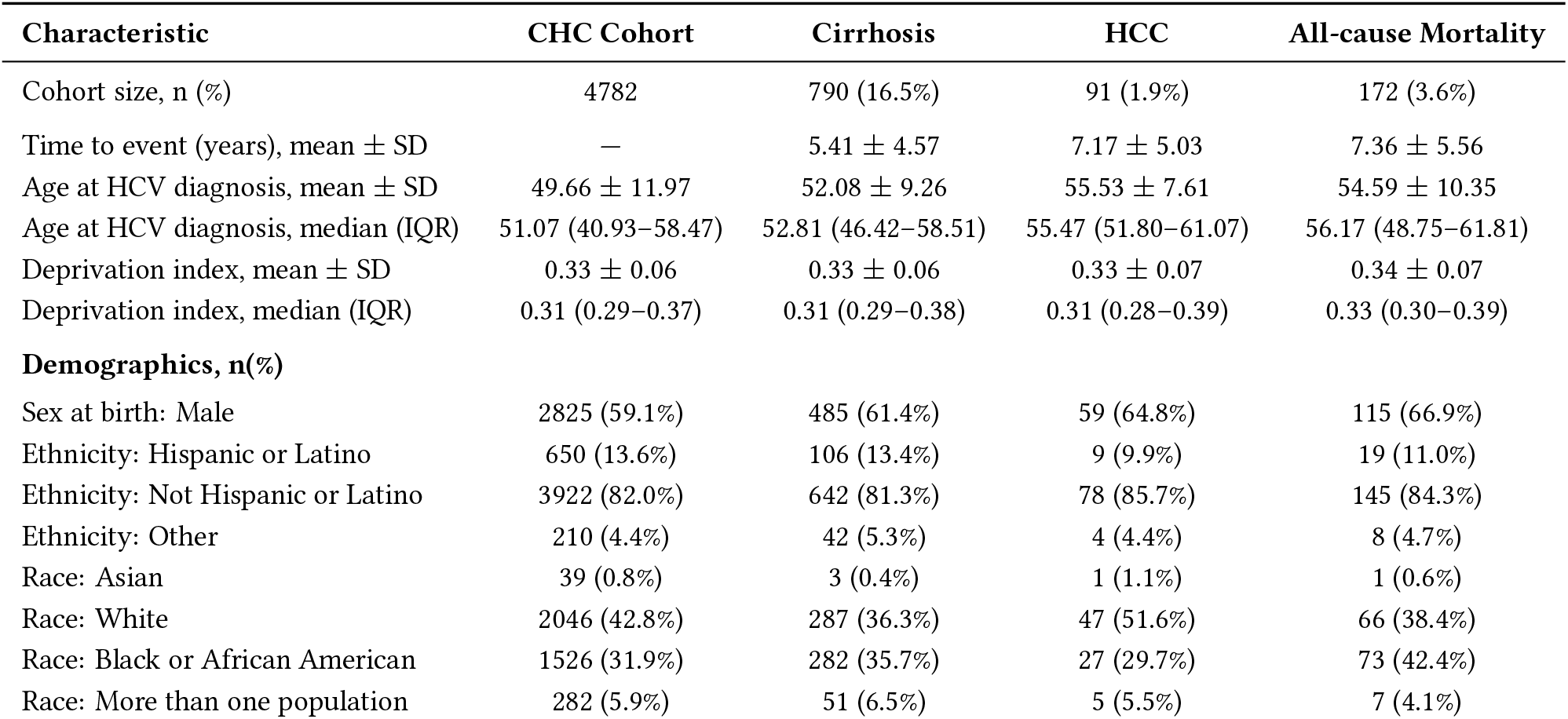
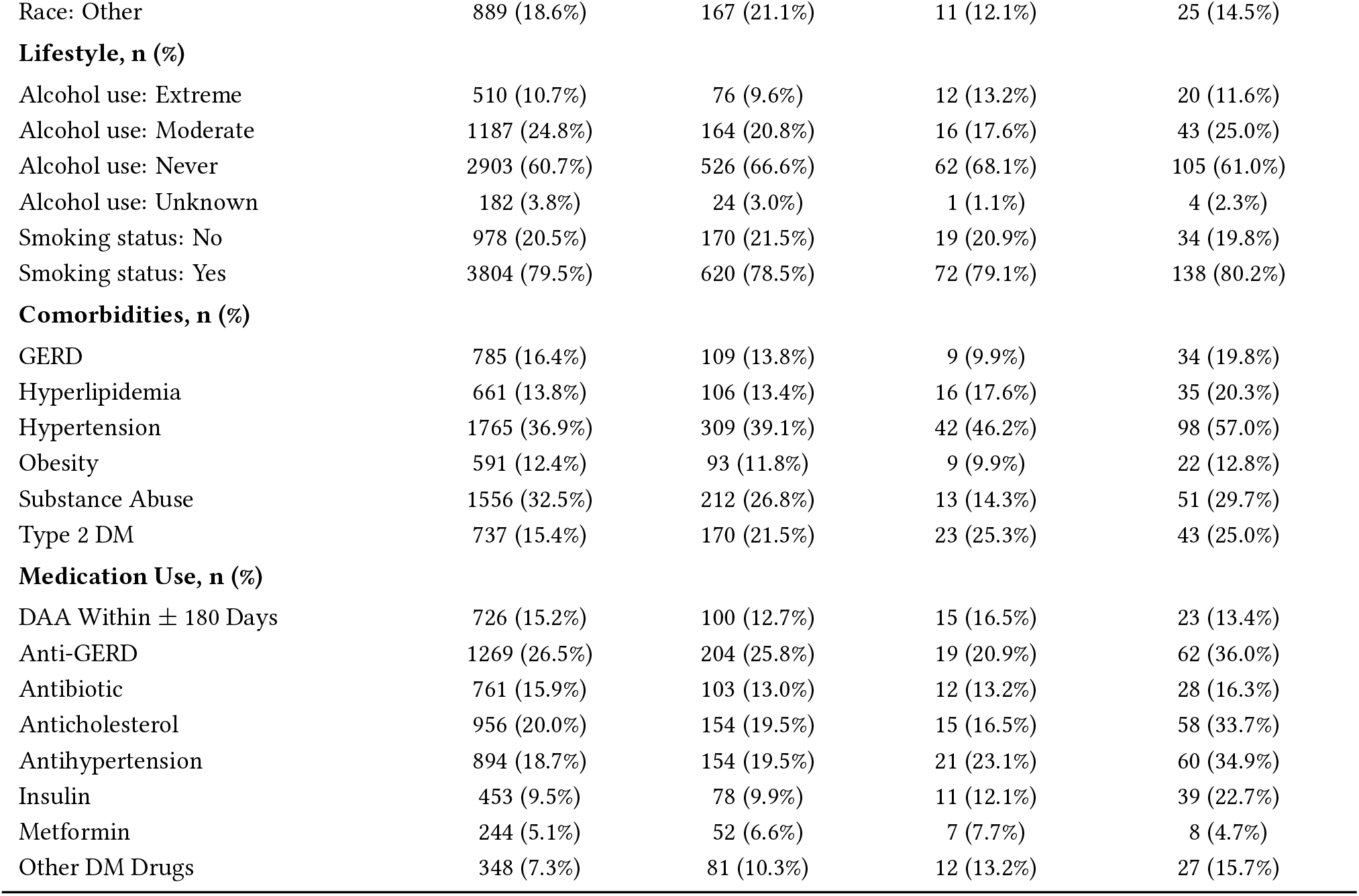
Baseline characteristics of the chronic hepatitis C (CHC) cohort overall and by incident outcomes. Values are reported as *n* (%) for categorical variables and as mean ±SD or median (IQR) for continuous variables.

| Characteristic | CHC Cohort | Cirrhosis | HCC | All-cause Mortality |
| --- | --- | --- | --- | --- |
| Cohort size, $n$ (%) | 4782 | 790 (16.5%) | 91 (1.9%) | 172 (3.6%) |
| Time to event (years), mean ± SD | — | 5.41 ± 4.57 | 7.17 ± 5.03 | 7.36 ± 5.56 |
| Age at HCV diagnosis, mean ± SD | 49.66 ± 11.97 | 52.08 ± 9.26 | 55.53 ± 7.61 | 54.59 ± 10.35 |
| Age at HCV diagnosis, median (IQR) | 51.07 (40.93–58.47) | 52.81 (46.42–58.51) | 55.47 (51.80–61.07) | 56.17 (48.75–61.81) |
| Deprivation index, mean ± SD | 0.33 ± 0.06 | 0.33 ± 0.06 | 0.33 ± 0.07 | 0.34 ± 0.07 |
| Deprivation index, median (IQR) | 0.31 (0.29–0.37) | 0.31 (0.29–0.38) | 0.31 (0.28–0.39) | 0.33 (0.30–0.39) |
| **Demographics, $n$(%)** |  |  |  |  |
| Sex at birth: Male | 2825 (59.1%) | 485 (61.4%) | 59 (64.8%) | 115 (66.9%) |
| Ethnicity: Hispanic or Latino | 650 (13.6%) | 106 (13.4%) | 9 (9.9%) | 19 (11.0%) |
| Ethnicity: Not Hispanic or Latino | 3922 (82.0%) | 642 (81.3%) | 78 (85.7%) | 145 (84.3%) |
| Ethnicity: Other | 210 (4.4%) | 42 (5.3%) | 4 (4.4%) | 8 (4.7%) |
| Race: Asian | 39 (0.8%) | 3 (0.4%) | 1 (1.1%) | 1 (0.6%) |
| Race: White | 2046 (42.8%) | 287 (36.3%) | 47 (51.6%) | 66 (38.4%) |
| Race: Black or African American | 1526 (31.9%) | 282 (35.7%) | 27 (29.7%) | 73 (42.4%) |
| Race: More than one population | 282 (5.9%) | 51 (6.5%) | 5 (5.5%) | 7 (4.1%) |
| Race: Other | 889 (18.6%) | 167 (21.1%) | 11 (12.1%) | 25 (14.5%) |
| **Lifestyle, n (%)** |  |  |  |  |
| Alcohol use: Extreme | 510 (10.7%) | 76 (9.6%) | 12 (13.2%) | 20 (11.6%) |
| Alcohol use: Moderate | 1187 (24.8%) | 164 (20.8%) | 16 (17.6%) | 43 (25.0%) |
| Alcohol use: Never | 2903 (60.7%) | 526 (66.6%) | 62 (68.1%) | 105 (61.0%) |
| Alcohol use: Unknown | 182 (3.8%) | 24 (3.0%) | 1 (1.1%) | 4 (2.3%) |
| Smoking status: No | 978 (20.5%) | 170 (21.5%) | 19 (20.9%) | 34 (19.8%) |
| Smoking status: Yes | 3804 (79.5%) | 620 (78.5%) | 72 (79.1%) | 138 (80.2%) |
| **Comorbidities, n (%)** |  |  |  |  |
| GERD | 785 (16.4%) | 109 (13.8%) | 9 (9.9%) | 34 (19.8%) |
| Hyperlipidemia | 661 (13.8%) | 106 (13.4%) | 16 (17.6%) | 35 (20.3%) |
| Hypertension | 1765 (36.9%) | 309 (39.1%) | 42 (46.2%) | 98 (57.0%) |
| Obesity | 591 (12.4%) | 93 (11.8%) | 9 (9.9%) | 22 (12.8%) |
| Substance Abuse | 1556 (32.5%) | 212 (26.8%) | 13 (14.3%) | 51 (29.7%) |
| Type 2 DM | 737 (15.4%) | 170 (21.5%) | 23 (25.3%) | 43 (25.0%) |
| **Medication Use, n (%)** |  |  |  |  |
| DAA Within ± 180 Days | 726 (15.2%) | 100 (12.7%) | 15 (16.5%) | 23 (13.4%) |
| Anti-GERD | 1269 (26.5%) | 204 (25.8%) | 19 (20.9%) | 62 (36.0%) |
| Antibiotic | 761 (15.9%) | 103 (13.0%) | 12 (13.2%) | 28 (16.3%) |
| Anticholesterol | 956 (20.0%) | 154 (19.5%) | 15 (16.5%) | 58 (33.7%) |
| Antihypertension | 894 (18.7%) | 154 (19.5%) | 21 (23.1%) | 60 (34.9%) |
| Insulin | 453 (9.5%) | 78 (9.9%) | 11 (12.1%) | 39 (22.7%) |
| Metformin | 244 (5.1%) | 52 (6.6%) | 7 (7.7%) | 8 (4.7%) |
| Other DM Drugs | 348 (7.3%) | 81 (10.3%) | 12 (13.2%) | 27 (15.7%) |

#### 3.3 Summary: Labs and vitals

**Supplementary Table 9:**
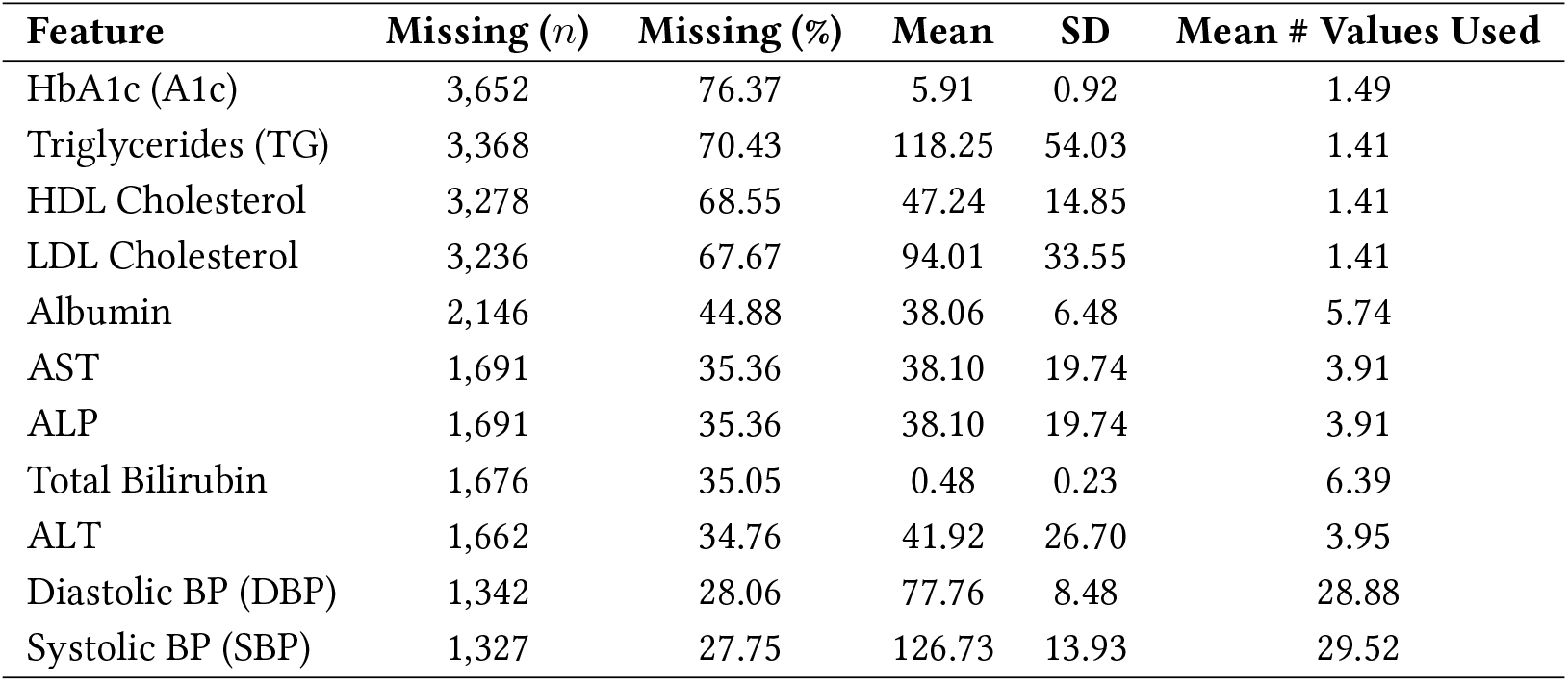
Summary of laboratory and vital sign features. For each feature, the number and percentage of missing values are reported, along with the mean and standard deviation computed among non-missing observations. The final column reports the mean number of observations per patient used to compute the aggregated mean value within the ±180-day window.

| Feature | Missing (*n*) | Missing (%) | Mean | SD | Mean # Values Used |
| --- | --- | --- | --- | --- | --- |
| HbA1c (A1c) | 3,652 | 76.37 | 5.91 | 0.92 | 1.49 |
| Triglycerides (TG) | 3,368 | 70.43 | 118.25 | 54.03 | 1.41 |
| HDL Cholesterol | 3,278 | 68.55 | 47.24 | 14.85 | 1.41 |
| LDL Cholesterol | 3,236 | 67.67 | 94.01 | 33.55 | 1.41 |
| Albumin | 2,146 | 44.88 | 38.06 | 6.48 | 5.74 |
| AST | 1,691 | 35.36 | 38.10 | 19.74 | 3.91 |
| ALP | 1,691 | 35.36 | 38.10 | 19.74 | 3.91 |
| Total Bilirubin | 1,676 | 35.05 | 0.48 | 0.23 | 6.39 |
| ALT | 1,662 | 34.76 | 41.92 | 26.70 | 3.95 |
| Diastolic BP (DBP) | 1,342 | 28.06 | 77.76 | 8.48 | 28.88 |
| Systolic BP (SBP) | 1,327 | 27.75 | 126.73 | 13.93 | 29.52 |

#### 3.4 Summary: Genomic features

**Supplementary Table 10:**
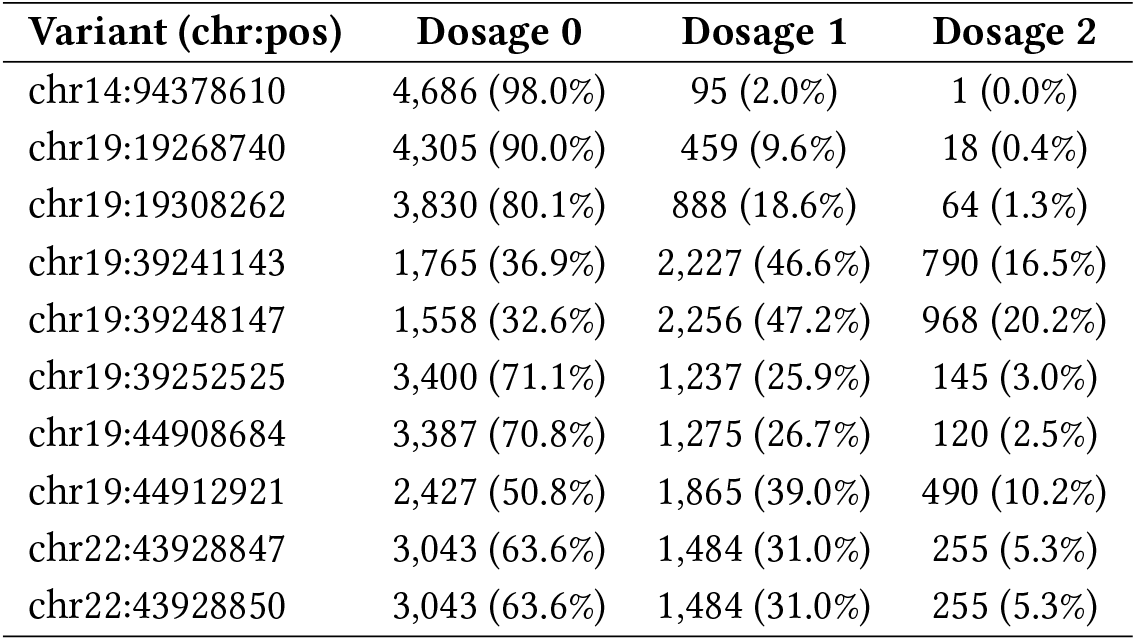
Distribution of genotype dosage levels for selected genetic variants. Counts and percentages are shown for each dosage category (0, 1, 2), calculated among participants with non-missing genotype data for the given locus.

| Variant (chr:pos) | Dosage 0 | Dosage 1 | Dosage 2 |
| --- | --- | --- | --- |
| chr14:94378610 | 4,686 (98.0%) | 95 (2.0%) | 1 (0.0%) |
| chr19:19268740 | 4,305 (90.0%) | 459 (9.6%) | 18 (0.4%) |
| chr19:19308262 | 3,830 (80.1%) | 888 (18.6%) | 64 (1.3%) |
| chr19:39241143 | 1,765 (36.9%) | 2,227 (46.6%) | 790 (16.5%) |
| chr19:39248147 | 1,558 (32.6%) | 2,256 (47.2%) | 968 (20.2%) |
| chr19:39252525 | 3,400 (71.1%) | 1,237 (25.9%) | 145 (3.0%) |
| chr19:44908684 | 3,387 (70.8%) | 1,275 (26.7%) | 120 (2.5%) |
| chr19:44912921 | 2,427 (50.8%) | 1,865 (39.0%) | 490 (10.2%) |
| chr22:43928847 | 3,043 (63.6%) | 1,484 (31.0%) | 255 (5.3%) |
| chr22:43928850 | 3,043 (63.6%) | 1,484 (31.0%) | 255 (5.3%) |

### 4 Model comparison

#### 4.1 Train set vs. held-out test

**Supplementary Table 11:** Comparison of training and test cohort characteristics across outcomes. Continuous variables are reported as mean ± SD. Categorical variables are reported as percentages within each split.

| Characteristic | Cirrhosis |  | HCC |  | All-cause Mortality |  |
| --- | --- | --- | --- | --- | --- | --- |
|  | Train (3824) | Test (956) | Train (3824) | Test (956) | Train (3498) | Test (874) |
| n of +cases (%) | 632 (16.5%) | 158 (16.5%) | 73 (1.9%) | 17 (1.8%) | 138 (3.9%) | 34 (3.9%) |
| Time to event (yrs) | 6.90 ± 5.55 | 6.45 ± 5.38 | 7.82 ± 5.96 | 7.65 ± 5.76 | 8.57 ± 5.84 | 8.39 ± 5.82 |
| Age at HCV diagnosis | 49.56 ± 11.93 | 50.05 ± 12.15 | 49.66 ± 11.90 | 49.64 ± 12.25 | 49.47 ± 11.84 | 49.68 ± 11.69 |
| Male (%) | 58.9 | 59.7 | 59.6 | 56.8 | 59.4 | 57.6 |
| Female (%) | 41.1 | 40.3 | 40.4 | 43.2 | 40.6 | 42.4 |
| **Race (%)** |  |  |  |  |  |  |
| White | 42.7 | 43.3 | 42.9 | 42.3 | 43.0 | 42.4 |
| Black/African American | 31.5 | 33.5 | 32.3 | 30.2 | 32.4 | 32.7 |
| Other | 18.9 | 17.5 | 18.3 | 19.9 | 17.5 | 20.6 |
| >1 population | 6.0 | 5.3 | 5.6 | 7.0 | 6.3 | 3.7 |
| Asian | 0.9 | 0.4 | 0.8 | 0.6 | 0.9 | 0.6 |
| **Ethnicity (%)** |  |  |  |  |  |  |
| Not Hispanic or Latino | 81.9 | 82.4 | 82.2 | 81.1 | 83.2 | 79.5 |
| Hispanic or Latino | 13.8 | 12.7 | 13.4 | 14.2 | 12.5 | 15.9 |
| Other | 4.3 | 4.9 | 4.3 | 4.7 | 4.3 | 4.6 |

#### 4.2 Hyperparameter tuning grid

**Supplementary Table 12:** Hyperparameter search space used for model tuning. Grid search was performed within the training data using cross-validation. Final hyperparameters were selected based on the highest cross-validated concordance index on the training set.

| Model | Hyperparameter | Search Values |
| --- | --- | --- |
| Coxnet (EN / LASSO) | alpha_min_ratio | {0.01, 0.05} |
|  | max_iter | {20000} |
| Random Survival Forest (RSF) | n_estimators | {500} |
|  | min_samples_leaf | {3, 5, 10} |
|  | min_samples_split | {10} |
|  | max_features | {sqrt} |
| Gradient Boosted Survival (GBSA) | learning_rate | {0.03, 0.05} |
|  | n_estimators | {300, 500} |
|  | max_depth | {2, 3} |
| Cox Neural Network (CoxNN) | learning rate (lr) | {1e-3} |
|  | epochs | {250, 300} |
|  | patience (early stopping) | {25} |
|  | weight decay (wd) | {1e-4, 5e-4} |
|  | dropout (pdrop) | {0.05, 0.10} |

#### 4.3 Model performance comparison

**Supplementary Table 13:** Model discrimination for predicting time to cirrhosis. C-index values are summarized as mean ± SD across cross-validation folds (training) and corresponding held-out test performance.

| Outcome | Feature set | Model | Train (CV) C-index | Test C-index |
| --- | --- | --- | --- | --- |
| Cirrhosis | Full | Coxnet-EN | 0.6705 ± 0.0223 | 0.6703 ± 0.0232 |
| | Full | Coxnet-LASSO | 0.6710 ± 0.0224 | 0.6709 ± 0.0231 |
| | Full | CoxNN | 0.6391 ± 0.0195 | 0.6477 ± 0.0254 |
| | Full | GBSA | 0.6604 ± 0.0191 | 0.6684 ± 0.0229 |
| | Full | RSF | 0.6629 ± 0.0186 | 0.6704 ± 0.0234 |
| | Top50% | Coxnet-EN | 0.6836 ± 0.0175 | 0.6684 ± 0.0237 |
| | Top50% | Coxnet-LASSO | 0.6830 ± 0.0183 | 0.6694 ± 0.0236 |
| | Top50% | CoxNN | 0.6731 ± 0.0166 | 0.6469 ± 0.0252 |
| | Top50% | GBSA | 0.6630 ± 0.0185 | 0.6658 ± 0.0231 |
| | Top50% | RSF | 0.6763 ± 0.0224 | 0.6548 ± 0.0242 |
| | Top25% | Coxnet-EN | 0.6843 ± 0.0131 | 0.6633 ± 0.0237 |
| | Top25% | Coxnet-LASSO | 0.6845 ± 0.0149 | 0.6620 ± 0.0237 |
| | Top25% | CoxNN | 0.6732 ± 0.0087 | 0.6440 ± 0.0258 |
| | Top25% | GBSA | 0.6630 ± 0.0173 | 0.6628 ± 0.0230 |
| | Top25% | RSF | 0.6681 ± 0.0218 | 0.6616 ± 0.0230 |

**Supplementary Table 14:** Time-dependent AUCs for predicting time to cirrhosis at 3, 5, and 10 years for training (CV) and held-out test sets.

| Outcome | Feature set | Model | Train (CV) AUC |  |  | Test AUC |  |  |
| --- | --- | --- | --- | --- | --- | --- | --- | --- |
|  |  |  | @3y | @5y | @10y | @3y | @5y | @10y |
| Cirrhosis | Full | Coxnet-EN | 0.6758 | 0.6938 | 0.7058 | 0.6920 | 0.6798 | 0.6851 |
|  | Full | Coxnet-LASSO | 0.6763 | 0.6941 | 0.7068 | 0.6942 | 0.6817 | 0.6838 |
|  | Full | CoxNN | 0.6362 | 0.6633 | 0.6647 | 0.6601 | 0.6566 | 0.6683 |
|  | Full | GBSA | 0.6657 | 0.6787 | 0.6842 | 0.6844 | 0.6600 | 0.6881 |
|  | Full | RSF | 0.6700 | 0.6861 | 0.6860 | 0.6743 | 0.6744 | 0.6947 |
|  | Top50% | Coxnet-EN | 0.6952 | 0.7059 | 0.7219 | 0.6922 | 0.6793 | 0.6835 |
|  | Top50% | Coxnet-LASSO | 0.6935 | 0.7053 | 0.7218 | 0.6953 | 0.6815 | 0.6822 |
|  | Top50% | CoxNN | 0.6802 | 0.6988 | 0.6988 | 0.6639 | 0.6546 | 0.6677 |
|  | Top50% | GBSA | 0.6696 | 0.6818 | 0.6886 | 0.6805 | 0.6571 | 0.6871 |
|  | Top50% | RSF | 0.6857 | 0.7011 | 0.7005 | 0.6640 | 0.6630 | 0.6722 |
|  | Top25% | Coxnet-EN | 0.6953 | 0.7072 | 0.7346 | 0.6908 | 0.6770 | 0.6910 |
|  | Top25% | Coxnet-LASSO | 0.6953 | 0.7070 | 0.7350 | 0.6895 | 0.6737 | 0.6865 |
|  | Top25% | CoxNN | 0.6798 | 0.7009 | 0.7098 | 0.6697 | 0.6482 | 0.6568 |
|  | Top25% | GBSA | 0.6662 | 0.6813 | 0.6925 | 0.6795 | 0.6573 | 0.6837 |
|  | Top25% | RSF | 0.6768 | 0.6885 | 0.6959 | 0.6657 | 0.6575 | 0.6770 |

**Supplementary Table 15:**
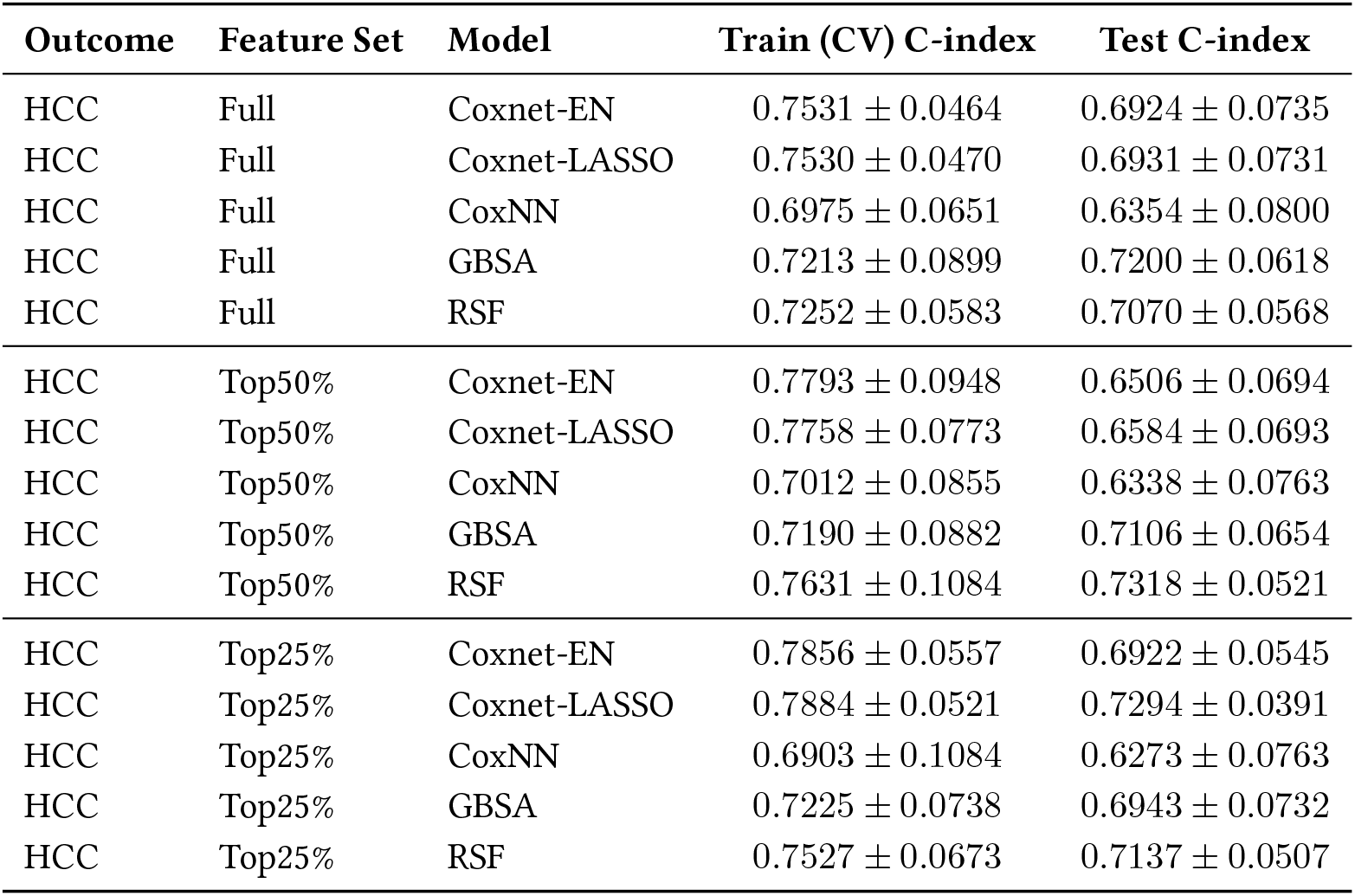
C-index performance for predicting hepatocellular carcinoma (HCC). Training results are summarized as mean ± SD across cross-validation folds (CV); test results are from the held-out dataset.

**Supplementary Table 16:** Time-dependent AUC performance for predicting hepatocellular carcinoma (HCC) at 3, 5, and 10 years. Training results are from cross-validation (CV); test results are from the held-out dataset.

| Outcome | Feature Set | Model | Train (CV) AUC |  |  | Test AUC |  |  |
| --- | --- | --- | --- | --- | --- | --- | --- | --- |
|  |  |  | @3y | @5y | @10y | @3y | @5y | @10y |
| HCC | Full | Coxnet-EN | 0.8268 | 0.7926 | 0.7380 | 0.7650 | 0.6803 | 0.6511 |
| HCC | Full | Coxnet-LASSO | 0.8337 | 0.7955 | 0.7332 | 0.7709 | 0.6847 | 0.6426 |
| HCC | Full | CoxNN | 0.7075 | 0.7089 | 0.6932 | 0.6479 | 0.5995 | 0.6294 |
| HCC | Full | GBSA | 0.7898 | 0.7524 | 0.7286 | 0.6403 | 0.7174 | 0.7172 |
| HCC | Full | RSF | 0.8319 | 0.7881 | 0.7136 | 0.6964 | 0.7015 | 0.7153 |
| HCC | Top50% | Coxnet-EN | 0.8862 | 0.7891 | 0.7901 | 0.6991 | 0.6262 | 0.6663 |
| HCC | Top50% | Coxnet-LASSO | 0.8699 | 0.7939 | 0.7888 | 0.7105 | 0.6415 | 0.6653 |
| HCC | Top50% | CoxNN | 0.7962 | 0.7021 | 0.7267 | 0.6854 | 0.5812 | 0.6468 |
| HCC | Top50% | GBSA | 0.7802 | 0.7561 | 0.7397 | 0.6280 | 0.7027 | 0.7477 |
| HCC | Top50% | RSF | 0.8383 | 0.8025 | 0.7665 | 0.6771 | 0.7322 | 0.7388 |
| HCC | Top25% | Coxnet-EN | 0.8623 | 0.8095 | 0.7976 | 0.6764 | 0.6909 | 0.7073 |
| HCC | Top25% | Coxnet-LASSO | 0.8571 | 0.8233 | 0.7956 | 0.7410 | 0.7399 | 0.7324 |
| HCC | Top25% | CoxNN | 0.7732 | 0.7037 | 0.7145 | 0.6523 | 0.5975 | 0.6230 |
| HCC | Top25% | GBSA | 0.7629 | 0.7641 | 0.7445 | 0.5990 | 0.6821 | 0.7345 |
| HCC | Top25% | RSF | 0.8483 | 0.8059 | 0.7482 | 0.6641 | 0.7131 | 0.7017 |

**Supplementary Table 17:** C-index performance for predicting all-cause mortality. Training results are summarized as mean ± SD across cross-validation folds (CV); test results are from the held-out dataset.

| Outcome | Feature Set | Model | Train (CV) C-index | Test C-index |
| --- | --- | --- | --- | --- |
| All-cause Mortality | Full | Coxnet-EN | 0.7165 ± 0.0450 | 0.7316 ± 0.0485 |
| All-cause Mortality | Full | Coxnet-LASSO | 0.7158 ± 0.0469 | 0.7279 ± 0.0520 |
| All-cause Mortality | Full | CoxNN | 0.6685 ± 0.0428 | 0.7128 ± 0.0455 |
| All-cause Mortality | Full | GBSA | 0.7240 ± 0.0592 | 0.7077 ± 0.0454 |
| All-cause Mortality | Full | RSF | 0.7595 ± 0.0463 | 0.7542 ± 0.0461 |
| All-cause Mortality | Top50% | Coxnet-EN | 0.7412 ± 0.0363 | 0.7442 ± 0.0480 |
| All-cause Mortality | Top50% | Coxnet-LASSO | 0.7415 ± 0.0413 | 0.7378 ± 0.0488 |
| All-cause Mortality | Top50% | CoxNN | 0.7246 ± 0.0197 | 0.6905 ± 0.0485 |
| All-cause Mortality | Top50% | GBSA | 0.7254 ± 0.0609 | 0.7051 ± 0.0483 |
| All-cause Mortality | Top50% | RSF | 0.7769 ± 0.0473 | 0.7481 ± 0.0438 |
| All-cause Mortality | Top25% | Coxnet-EN | 0.7506 ± 0.0404 | 0.7087 ± 0.0525 |
| All-cause Mortality | Top25% | Coxnet-LASSO | 0.7453 ± 0.0444 | 0.7019 ± 0.0544 |
| All-cause Mortality | Top25% | CoxNN | 0.7059 ± 0.0647 | 0.6847 ± 0.0515 |
| All-cause Mortality | Top25% | GBSA | 0.7295 ± 0.0624 | 0.7103 ± 0.0469 |
| All-cause Mortality | Top25% | RSF | 0.7780 ± 0.0415 | 0.7393 ± 0.0402 |

**Supplementary Table 18:** Time-dependent AUC performance for predicting all-cause mortality at 3, 5, and 10 years. Training results are from cross-validation (CV); test results are from the held-out dataset.

| Outcome | Feature Set | Model | Train (CV) AUC |  |  | Test AUC |  |  |
| --- | --- | --- | --- | --- | --- | --- | --- | --- |
|  |  |  | @3y | @5y | @10y | @3y | @5y | @10y |
| All-cause Mortality | Full | Coxnet-EN | 0.7254 | 0.7519 | 0.7540 | 0.7119 | 0.7586 | 0.7822 |
| All-cause Mortality | Full | Coxnet-LASSO | 0.7198 | 0.7458 | 0.7561 | 0.7011 | 0.7531 | 0.7783 |
| All-cause Mortality | Full | CoxNN | 0.6796 | 0.7006 | 0.6840 | 0.6462 | 0.7372 | 0.7652 |
| All-cause Mortality | Full | GBSA | 0.7172 | 0.7287 | 0.7816 | 0.7228 | 0.7508 | 0.7244 |
| All-cause Mortality | Full | RSF | 0.7570 | 0.8002 | 0.8038 | 0.8066 | 0.8080 | 0.7851 |
| All-cause Mortality | Top50% | Coxnet-EN | 0.7617 | 0.7619 | 0.7697 | 0.7354 | 0.7734 | 0.7928 |
| All-cause Mortality | Top50% | Coxnet-LASSO | 0.7741 | 0.7651 | 0.7710 | 0.7210 | 0.7628 | 0.7913 |
| All-cause Mortality | Top50% | CoxNN | 0.7506 | 0.7638 | 0.7407 | 0.6978 | 0.7105 | 0.7274 |
| All-cause Mortality | Top50% | GBSA | 0.7237 | 0.7314 | 0.7835 | 0.7234 | 0.7468 | 0.7122 |
| All-cause Mortality | Top50% | RSF | 0.7890 | 0.8113 | 0.8268 | 0.7812 | 0.8053 | 0.7858 |
| All-cause Mortality | Top25% | Coxnet-EN | 0.7817 | 0.7793 | 0.7909 | 0.7165 | 0.7392 | 0.7287 |
| All-cause Mortality | Top25% | Coxnet-LASSO | 0.7761 | 0.7722 | 0.7843 | 0.6996 | 0.7335 | 0.7249 |
| All-cause Mortality | Top25% | CoxNN | 0.7284 | 0.7412 | 0.7322 | 0.6696 | 0.7089 | 0.7096 |
| All-cause Mortality | Top25% | GBSA | 0.7292 | 0.7356 | 0.7851 | 0.7319 | 0.7544 | 0.7242 |
| All-cause Mortality | Top25% | RSF | 0.7862 | 0.8124 | 0.8152 | 0.7789 | 0.7999 | 0.7605 |

#### 4.4 Handling missing data

**Supplementary Table 19:**
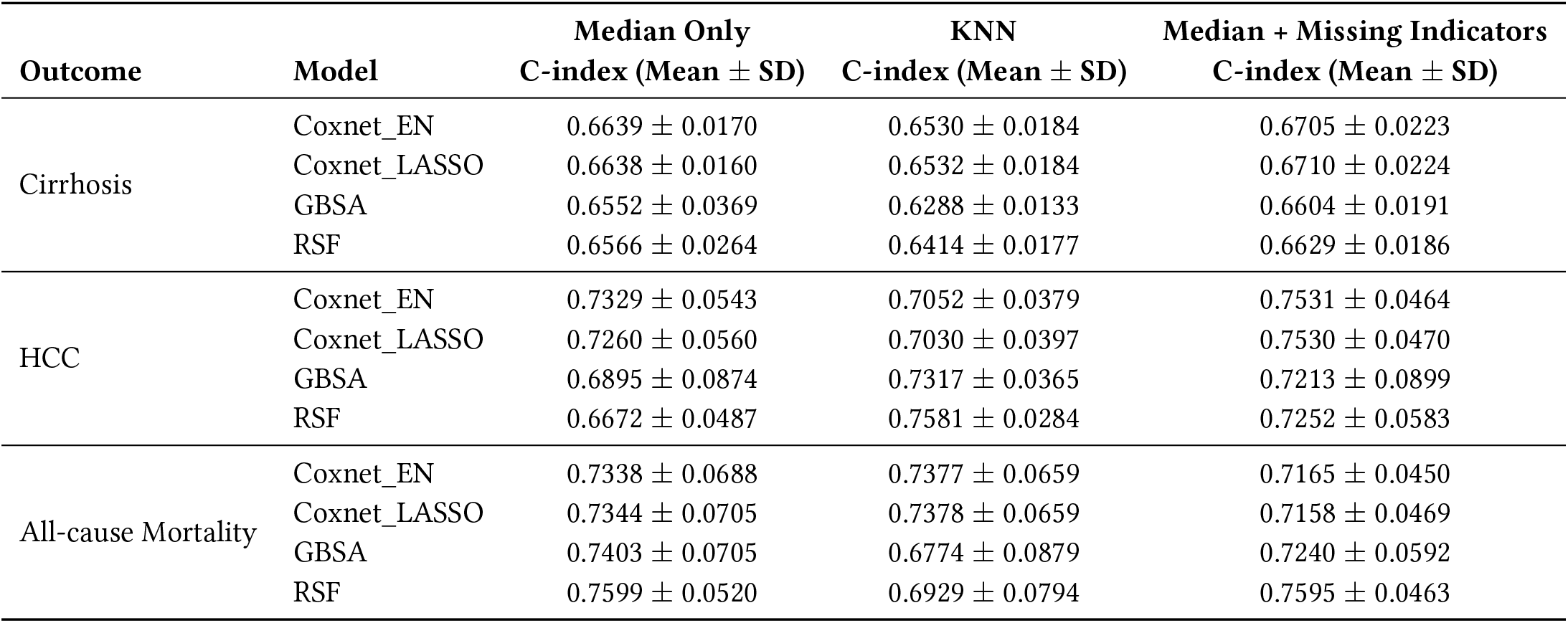
Comparison of model performance (C-index) under different imputation strategies. Values are reported as mean ± SD across cross-validation folds.

| Outcome | Model | Median Only | KNN | Median + Missing Indicators |
| --- | --- | --- | --- | --- |
| | | C-index (Mean ± SD) | C-index (Mean ± SD) | C-index (Mean ± SD) |
| Cirrhosis | Coxnet_EN | 0.6639 ± 0.0170 | 0.6530 ± 0.0184 | 0.6705 ± 0.0223 |
| | Coxnet_LASSO | 0.6638 ± 0.0160 | 0.6532 ± 0.0184 | 0.6710 ± 0.0224 |
| | GBSA | 0.6552 ± 0.0369 | 0.6288 ± 0.0133 | 0.6604 ± 0.0191 |
| | RSF | 0.6566 ± 0.0264 | 0.6414 ± 0.0177 | 0.6629 ± 0.0186 |
| HCC | Coxnet_EN | 0.7329 ± 0.0543 | 0.7052 ± 0.0379 | 0.7531 ± 0.0464 |
| | Coxnet_LASSO | 0.7260 ± 0.0560 | 0.7030 ± 0.0397 | 0.7530 ± 0.0470 |
| | GBSA | 0.6895 ± 0.0874 | 0.7317 ± 0.0365 | 0.7213 ± 0.0899 |
| | RSF | 0.6672 ± 0.0487 | 0.7581 ± 0.0284 | 0.7252 ± 0.0583 |
| All-cause Mortality | Coxnet_EN | 0.7338 ± 0.0688 | 0.7377 ± 0.0659 | 0.7165 ± 0.0450 |
| | Coxnet_LASSO | 0.7344 ± 0.0705 | 0.7378 ± 0.0659 | 0.7158 ± 0.0469 |
| | GBSA | 0.7403 ± 0.0705 | 0.6774 ± 0.0879 | 0.7240 ± 0.0592 |
| | RSF | 0.7599 ± 0.0520 | 0.6929 ± 0.0794 | 0.7595 ± 0.0463 |

### 5 Model evaluation and explainability

#### 5.1 Permutation importance for feature selection

**Supplementary Figure 2.**
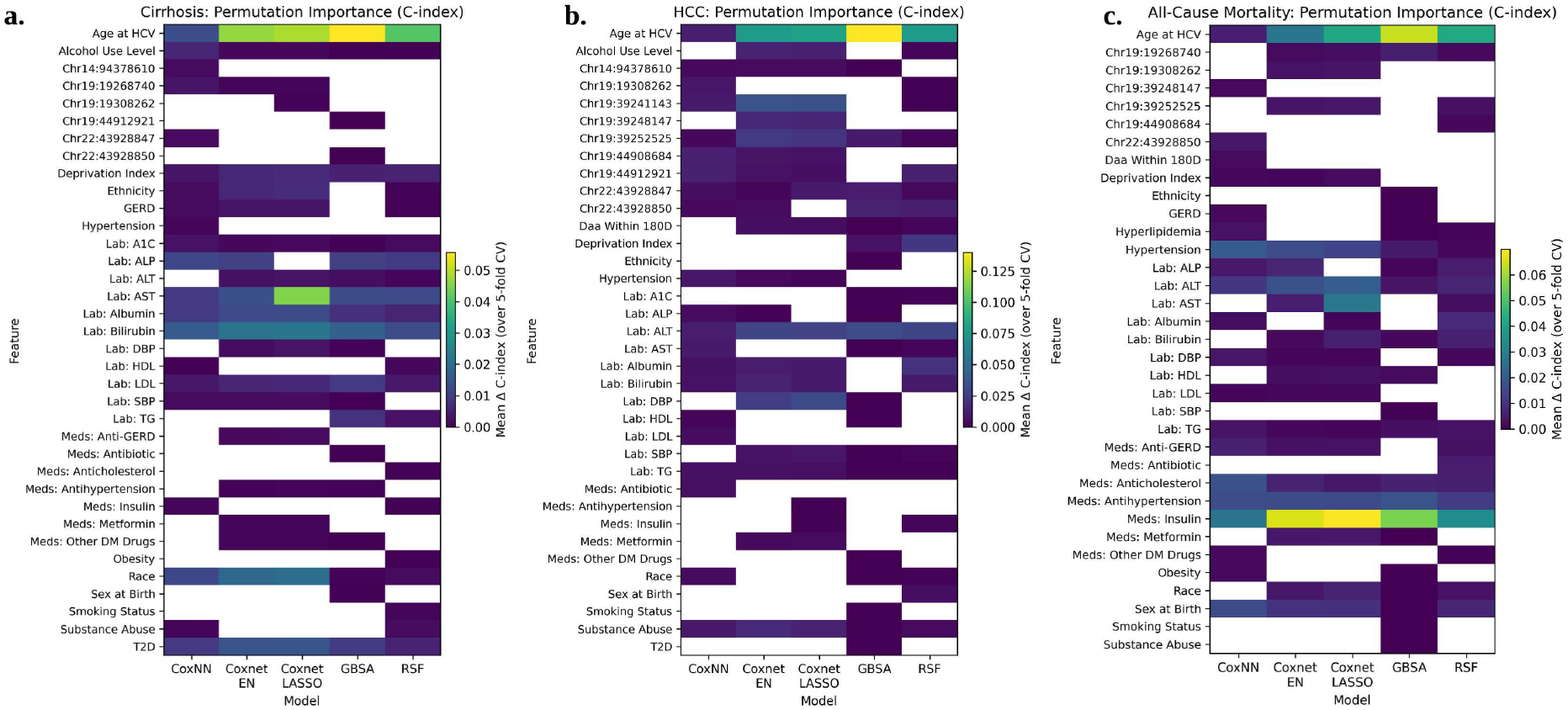
Permutation importance based on C-index across models and outcomes. Heatmaps display mean permutation importance scores (mean Δ C-index) computed from 5-fold cross-validation on the training data for each outcome: (**a**) cirrhosis, (**b**) hepatocellular carcinoma (HCC), and (**c**) all-cause mortality. For each feature, importance was quantified as the decrease in C-index after permutation, averaged across folds. Columns represent models (CoxNN, Coxnet-EN, Coxnet-LASSO, GBSA, and RSF), and rows represent candidate predictors. Higher values indicate greater contribution to predictive discrimination. Blank cells indicate that a feature was not selected within the corresponding reduced feature set. These cross-validated permutation importance estimates were subsequently used to derive reduced feature sets (Top50% and Top25%) for downstream model training and evaluation.

#### 5.2 KM-risk stratification

**Supplementary Figure 3.**
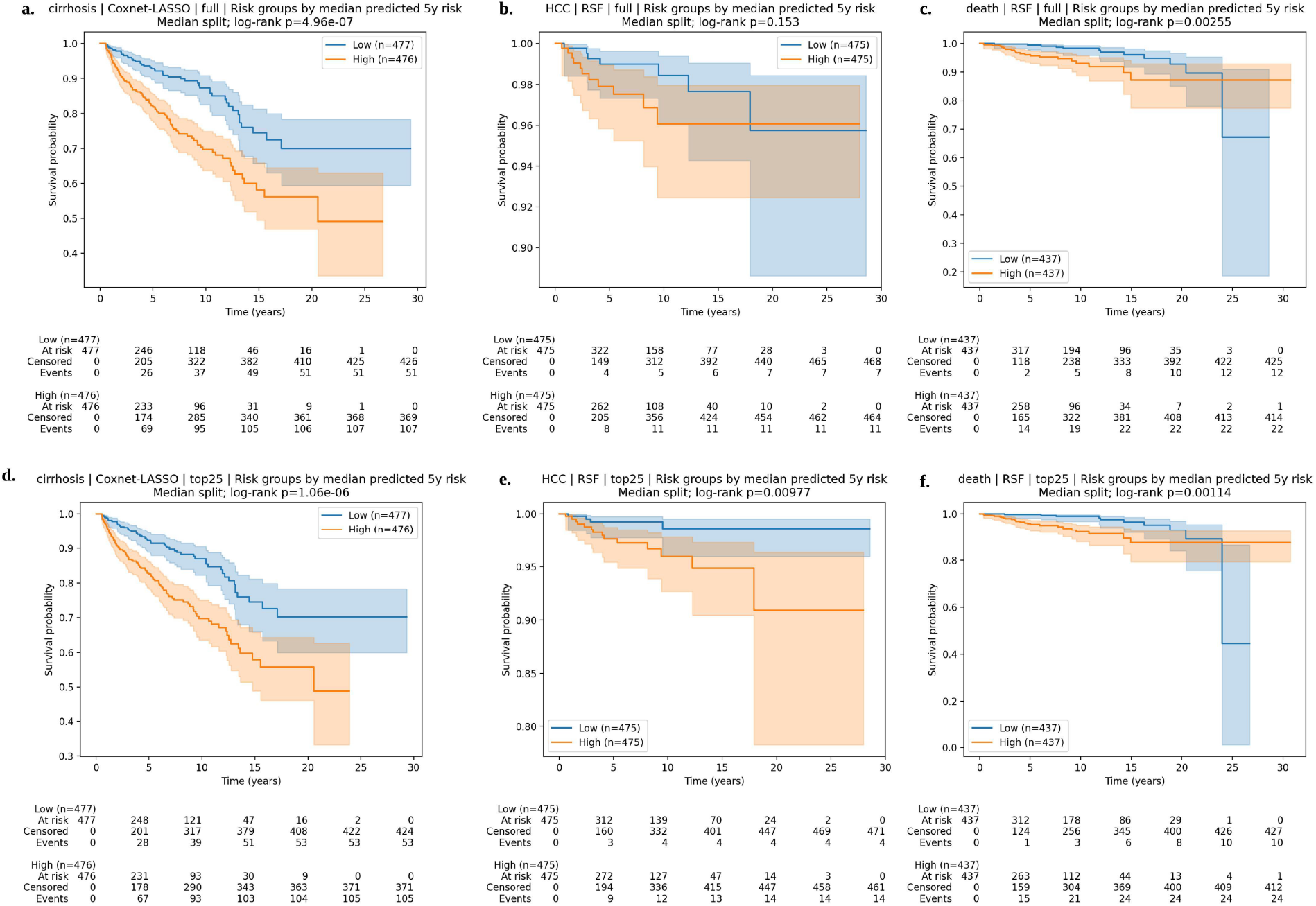
Kaplan–Meier survival curves stratified by median predicted 5-year risk across outcomes and feature sets. For each outcome, participants were stratified into high- and low-risk groups based on the median predicted 5-year risk derived from the respective survival model. Panels (**a–c**) show results using the full feature set, and panels (**d–f**) show results using the Top25% reduced feature set. (**a, d**) Cirrhosis predicted by Coxnet-LASSO; (**b, e**) hepatocellular carcinoma (HCC) predicted by RSF; (**c, f**) all-cause mortality predicted by RSF. Shaded regions represent 95% confidence intervals. Numbers at risk, cumulative events, and censoring counts are displayed below each panel. Log-rank p-values assess survival differences between risk groups. Significant separation between high- and low-risk groups demonstrates effective risk discrimination and clinical stratification performance.

#### 5.3 Calibration on held-out test

**Supplementary Table 20:**
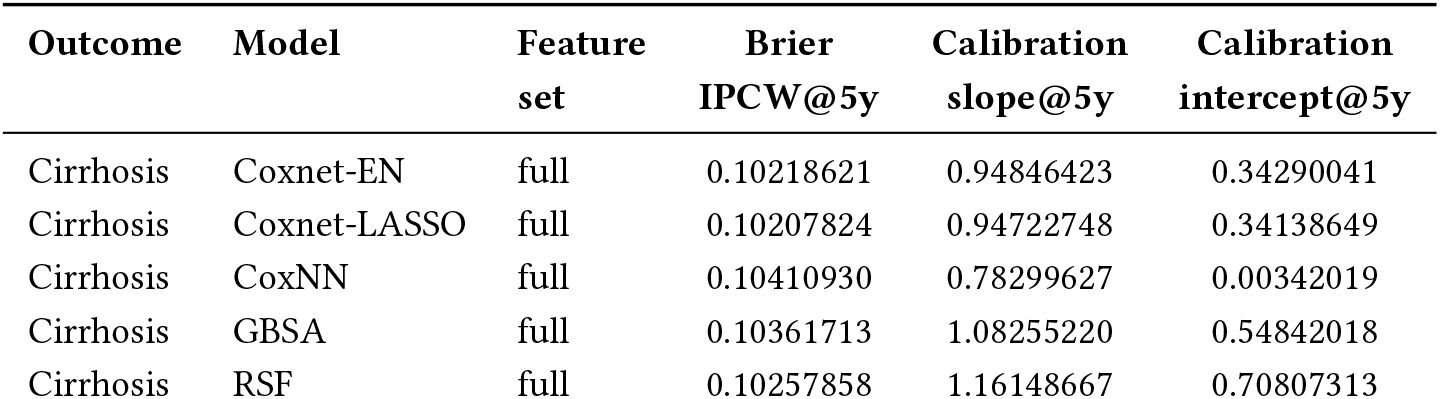
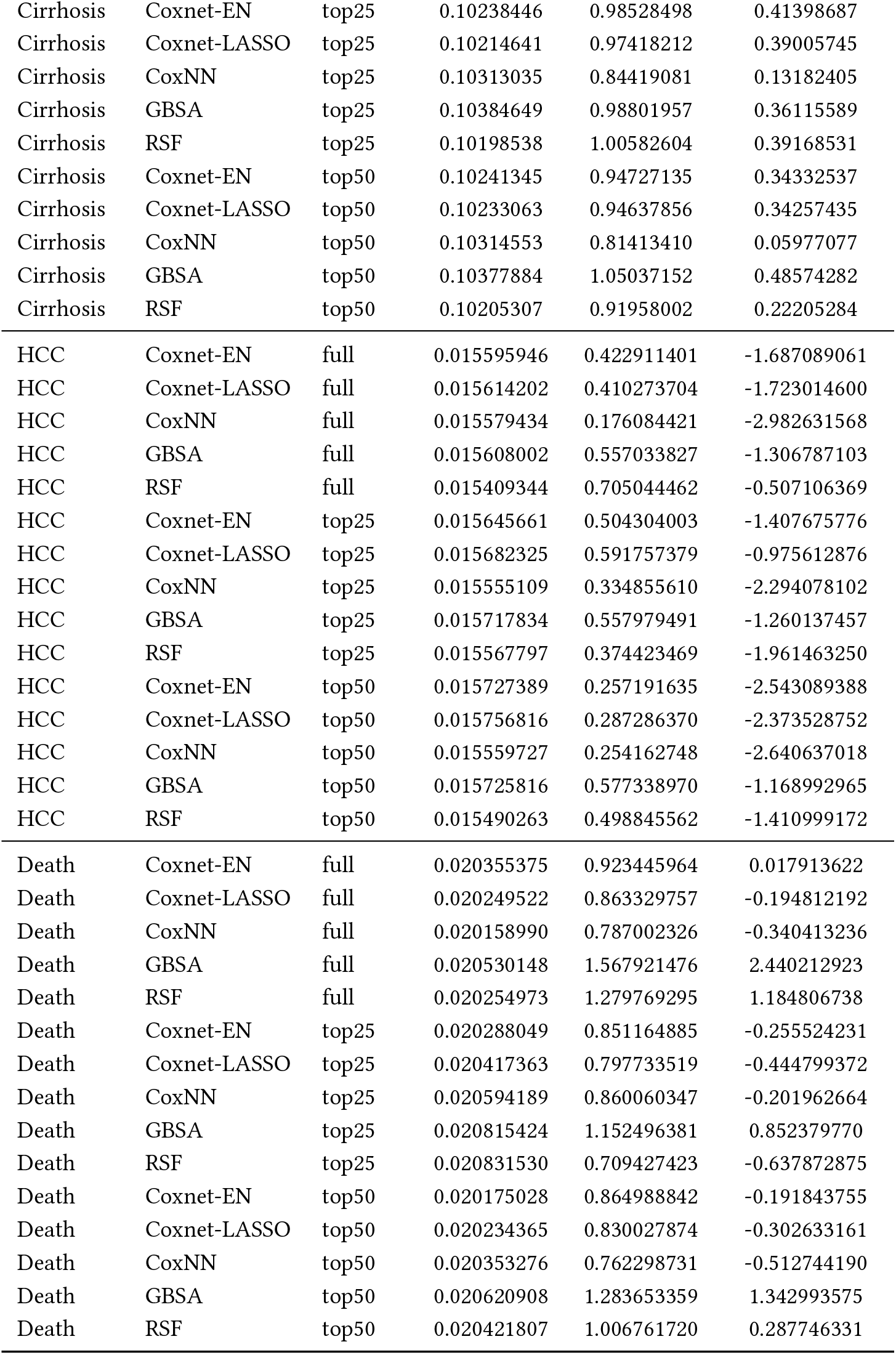
Model calibration and discrimination at 5 years across outcomes, models, and feature sets. Brier score is IPCW-adjusted. Calibration slope and intercept are computed at 5 years.

| Outcome | Model | Feature set | Brier IPCW@5y | Calibration slope@5y | Calibration intercept@5y |
| --- | --- | --- | --- | --- | --- |
| Cirrhosis | Coxnet-EN | full | 0.10218621 | 0.94846423 | 0.34290041 |
| Cirrhosis | Coxnet-LASSO | full | 0.10207824 | 0.94722748 | 0.34138649 |
| Cirrhosis | CoxNN | full | 0.10410930 | 0.78299627 | 0.00342019 |
| Cirrhosis | GBSA | full | 0.10361713 | 1.08255220 | 0.54842018 |
| Cirrhosis | RSF | full | 0.10257858 | 1.16148667 | 0.70807313 |
| Cirrhosis | Coxnet-EN | top25 | 0.10238446 | 0.98528498 | 0.41398687 |
| Cirrhosis | Coxnet-LASSO | top25 | 0.10214641 | 0.97418212 | 0.39005745 |
| Cirrhosis | CoxNN | top25 | 0.10313035 | 0.84419081 | 0.13182405 |
| Cirrhosis | GBSA | top25 | 0.10384649 | 0.98801957 | 0.36115589 |
| Cirrhosis | RSF | top25 | 0.10198538 | 1.00582604 | 0.39168531 |
| Cirrhosis | Coxnet-EN | top50 | 0.10241345 | 0.94727135 | 0.34332537 |
| Cirrhosis | Coxnet-LASSO | top50 | 0.10233063 | 0.94637856 | 0.34257435 |
| Cirrhosis | CoxNN | top50 | 0.10314553 | 0.81413410 | 0.05977077 |
| Cirrhosis | GBSA | top50 | 0.10377884 | 1.05037152 | 0.48574282 |
| Cirrhosis | RSF | top50 | 0.10205307 | 0.91958002 | 0.22205284 |
| HCC | Coxnet-EN | full | 0.015595946 | 0.422911401 | -1.687089061 |
| HCC | Coxnet-LASSO | full | 0.015614202 | 0.410273704 | -1.723014600 |
| HCC | CoxNN | full | 0.015579434 | 0.176084421 | -2.982631568 |
| HCC | GBSA | full | 0.015608002 | 0.557033827 | -1.306787103 |
| HCC | RSF | full | 0.015409344 | 0.705044462 | -0.507106369 |
| HCC | Coxnet-EN | top25 | 0.015645661 | 0.504304003 | -1.407675776 |
| HCC | Coxnet-LASSO | top25 | 0.015682325 | 0.591757379 | -0.975612876 |
| HCC | CoxNN | top25 | 0.015555109 | 0.334855610 | -2.294078102 |
| HCC | GBSA | top25 | 0.015717834 | 0.557979491 | -1.260137457 |
| HCC | RSF | top25 | 0.015567797 | 0.374423469 | -1.961463250 |
| HCC | Coxnet-EN | top50 | 0.015727389 | 0.257191635 | -2.543089388 |
| HCC | Coxnet-LASSO | top50 | 0.015756816 | 0.287286370 | -2.373528752 |
| HCC | CoxNN | top50 | 0.015559727 | 0.254162748 | -2.640637018 |
| HCC | GBSA | top50 | 0.015725816 | 0.577338970 | -1.168992965 |
| HCC | RSF | top50 | 0.015490263 | 0.498845562 | -1.410999172 |
| Death | Coxnet-EN | full | 0.020355375 | 0.923445964 | 0.017913622 |
| Death | Coxnet-LASSO | full | 0.020249522 | 0.863329757 | -0.194812192 |
| Death | CoxNN | full | 0.020158990 | 0.787002326 | -0.340413236 |
| Death | GBSA | full | 0.020530148 | 1.567921476 | 2.440212923 |
| Death | RSF | full | 0.020254973 | 1.279769295 | 1.184806738 |
| Death | Coxnet-EN | top25 | 0.020288049 | 0.851164885 | -0.255524231 |
| Death | Coxnet-LASSO | top25 | 0.020417363 | 0.797733519 | -0.444799372 |
| Death | CoxNN | top25 | 0.020594189 | 0.860060347 | -0.201962664 |
| Death | GBSA | top25 | 0.020815424 | 1.152496381 | 0.852379770 |
| Death | RSF | top25 | 0.020831530 | 0.709427423 | -0.637872875 |
| Death | Coxnet-EN | top50 | 0.020175028 | 0.864988842 | -0.191843755 |
| Death | Coxnet-LASSO | top50 | 0.020234365 | 0.830027874 | -0.302633161 |
| Death | CoxNN | top50 | 0.020353276 | 0.762298731 | -0.512744190 |
| Death | GBSA | top50 | 0.020620908 | 1.283653359 | 1.342993575 |
| Death | RSF | top50 | 0.020421807 | 1.006761720 | 0.287746331 |

#### 5.4 SHAP analysis on held-out test

**Supplementary Figure 4.**
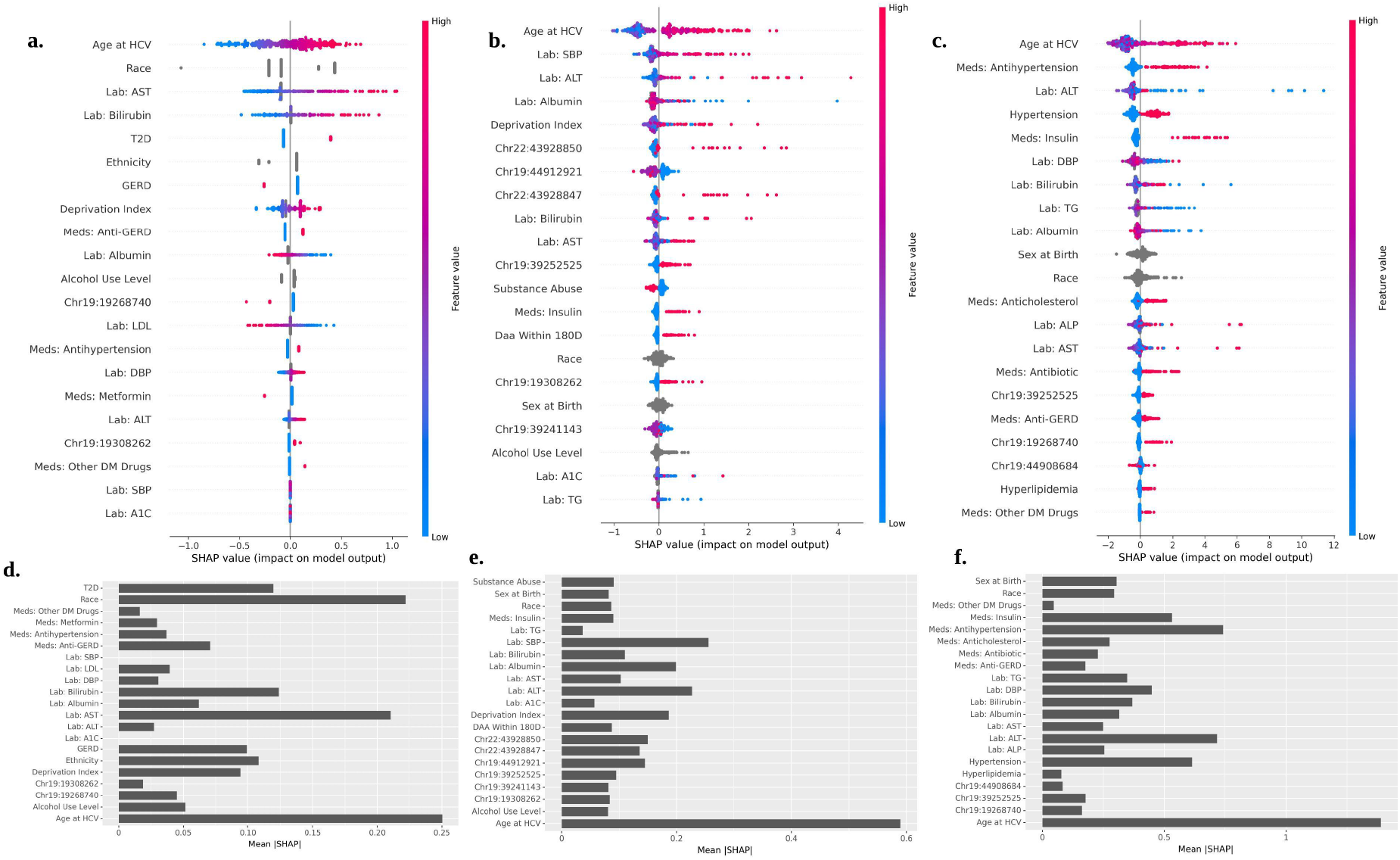
SHAP-based model interpretability for Top50% reduced feature sets across outcomes. Panels (**a–c**) show SHAP summary (beeswarm) plots illustrating the distribution of feature contributions to model predictions, computed on the training cross-validation folds. Each point represents an individual participant; the x-axis indicates the SHAP value (impact on model output), and color encodes feature value (low to high). Panels (**d–f**) display corresponding mean absolute SHAP values (*mean* |*SHAP*|), ranking features by overall importance.(**a, d**) Cirrhosis; (**b, e**) hepatocellular carcinoma (HCC); (**c, f**) all-cause mortality.

#### 5.5 Genomic and medication features contribution

**Supplementary Figure 5.**
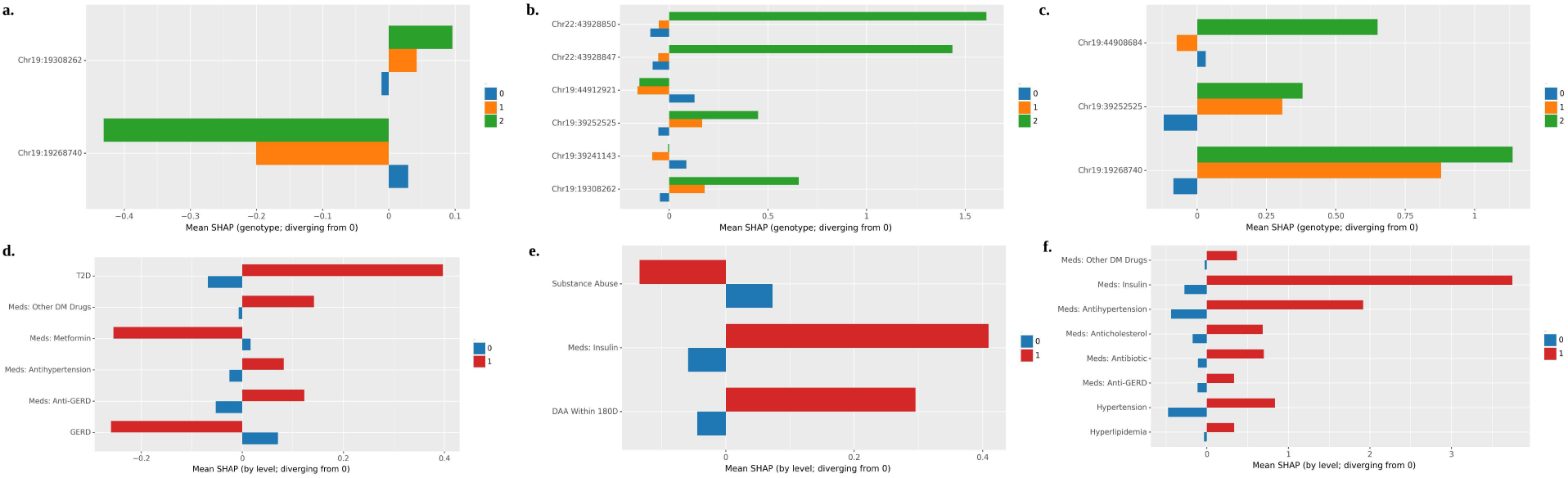
Genotype- and level-specific SHAP effects for selected features in Top50% reduced models. Panels (**a–c**) show mean SHAP values stratified by genotype dosage levels (0, 1, 2) for selected genetic variants, illustrating direction and magnitude of deviation from the reference (zero-impact baseline). Panels (**d–f**) present mean SHAP values stratified by binary clinical feature levels (0 vs 1), highlighting differential contribution of comorbidities and medication exposures to model predictions.(**a, d**) Cirrhosis; (**b, e**) hepatocellular carcinoma (HCC); (**c, f**) all-cause mortality. Positive SHAP values indicate increased predicted risk, whereas negative values indicate reduced predicted risk.

#### 5.6 Case study

**Supplementary Figure 6.**
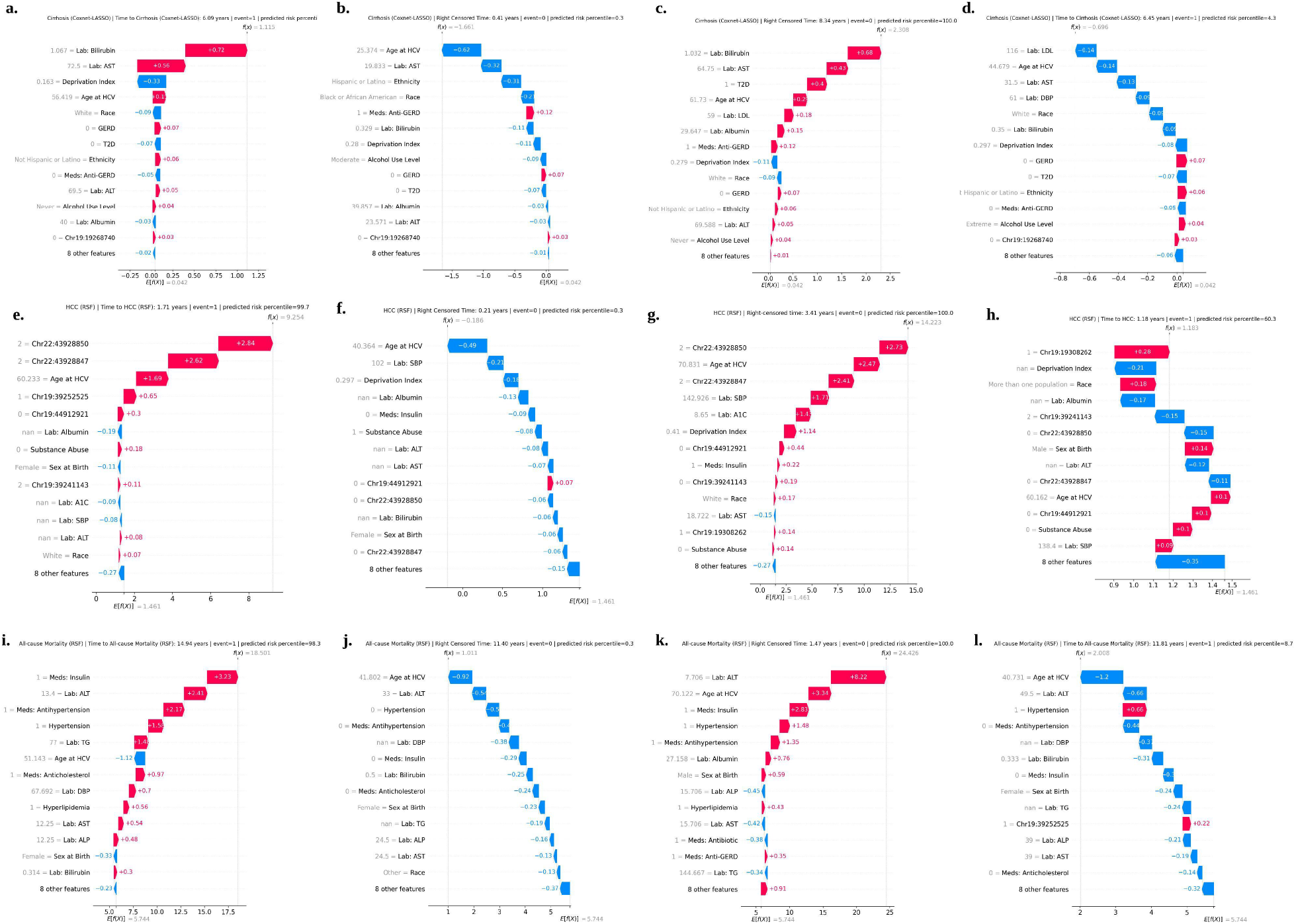
Representative individual-level SHAP waterfall plots across outcomes and prediction categories. Each panel displays local feature contributions (SHAP values) for selected individuals from the held-out test set. For each outcome, four representative cases are shown: true positive (TP), true negative (TN), false positive (FP), and false negative (FN), selected based on predicted risk percentiles. Panels (**a–d**) correspond to cirrhosis (Coxnet-LASSO model), (**e–h**) to hepatocellular carcinoma (HCC; RSF model), and (**i–l**) to all-cause mortality (RSF model). Horizontal bars indicate the direction and magnitude of each feature’s contribution to the individual’s predicted risk relative to the baseline expectation. Red bars increase predicted risk, while blue bars decrease predicted risk. The cumulative sum of feature effects explains the model’s final prediction for that individual.

#### 5.7 Competing risk estimates

**Supplementary Figure 7.**
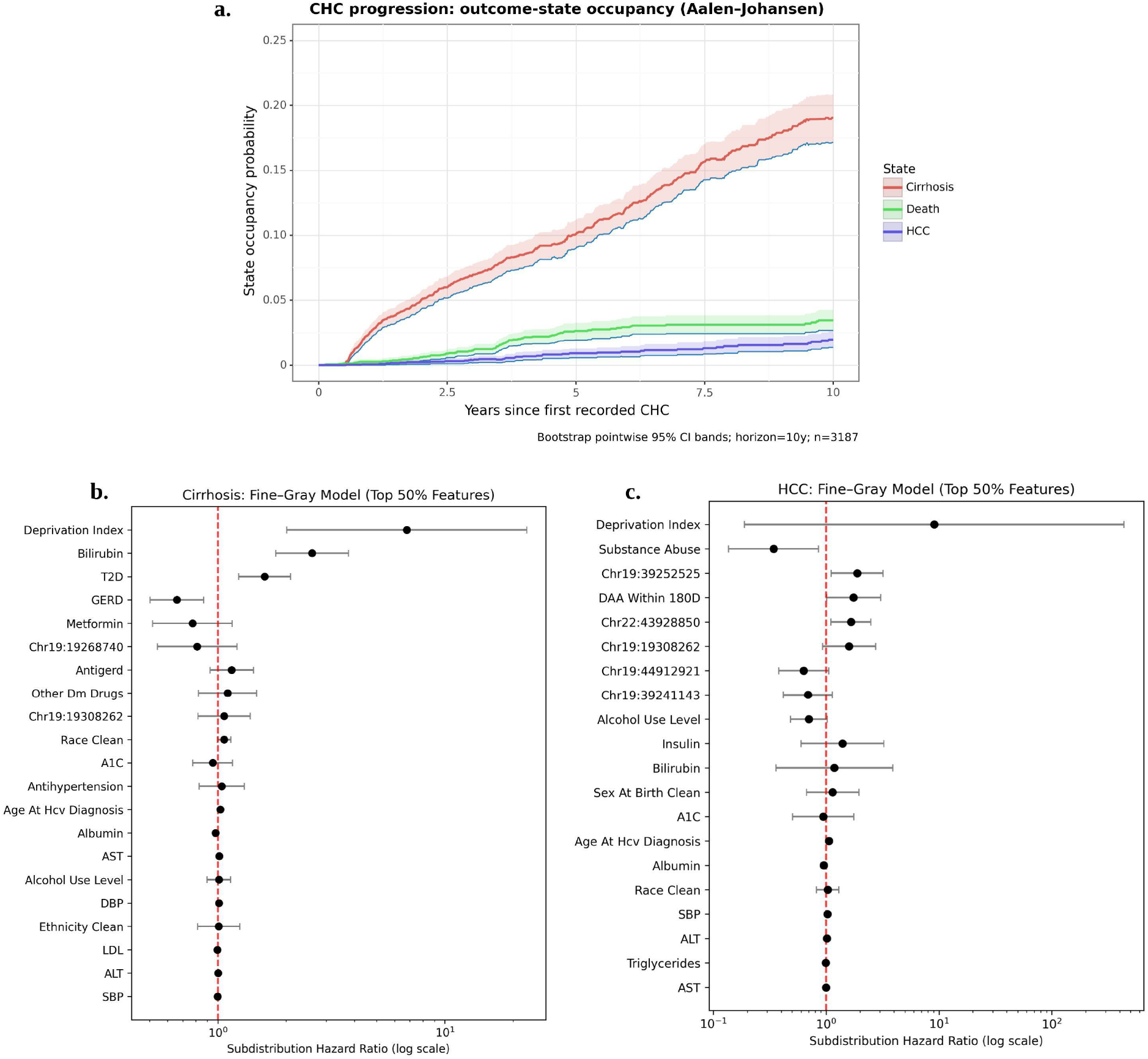
Multi-state CHC progression and competing-risk regression results. **(a)** Outcome-state occupancy probabilities estimated using the Aalen–Johansen estimator over a 10-year horizon following first recorded chronic hepatitis C (CHC). Curves represent the cumulative probability of occupying each state (cirrhosis, hepatocellular carcinoma [HCC], and death), with bootstrap pointwise 95% confidence intervals. Cirrhosis demonstrates the highest cumulative incidence over time, followed by death and HCC, reflecting the predominant intermediate disease transition pattern in this cohort. **(b)** Fine–Gray competing-risk regression model for cirrhosis using the top 50% of selected features. Points represent subdistribution hazard ratios (SHR) on a logarithmic scale, with horizontal lines indicating 95% confidence intervals. The red dashed vertical line denotes SHR = 1. Features to the right of 1 are associated with increased subdistribution hazard, while those to the left indicate protective associations. **(c)** Fine–Gray competing-risk regression model for HCC using the top 50% selected features, presented analogously. Deprivation index and selected clinical and genetic features demonstrate varying magnitudes of association, highlighting outcome-specific risk structures under competing risks.

## References

[1] World Health Organization. Global Hepatitis Report 2017. Tech. Rep., World Health Organization, Geneva, Switzerland (2017). https://www.who.int/publications/i/item/9789241565455.

[2] Leone, N. & Rizzetto, M. Natural history of hepatitis C virus infection: From chronic hepatitis to cirrhosis, to hepatocellular carcinoma. Minerva Gastroenterology and Dietology 51, 31–46 (2005).

[3] Devarbhavi, H. et al. Global burden of liver disease: 2023 update. Journal of Hepatology 79, 516–537 (2023).

[4] Luan, C.-H. et al. Residual risk of hepatocellular carcinoma development for chronic hepatitis C patients treated by all-oral direct-acting antivirals with sustained virological response. Journal of the Chinese Medical Association 86, 795–805 (2023).

[5] European Association for the Study of the Liver, Clinical Practice Guidelines Panel, EASL Governing Board Representative & Panel Members. EASL recommendations on treatment of hepatitis C: Final update of the series. Journal of Hepatology 73, 1170–1218 (2020).

[6] Bhattacharya, D., Aronsohn, A., Price, J. & Lo Re, V. Hepatitis C Guidance 2023 Update: AASLD-IDSA Recommendations for Testing, Managing, and Treating Hepatitis C Virus Infection. Clinical Infectious Diseases (2023).

[7] Ahumada, A., Rayón, L., Usón, C., Bañares, R. & Alonso Lopez, S. Hepatocellular carcinoma risk after viral response in hepatitis C virus-advanced fibrosis: Who to screen and for how long? World Journal of Gastroenterology 27, 6737–6749 (2021).

[8] Ioannou, G. N. et al. Development of models estimating the risk of hepatocellular carcinoma after antiviral treatment for hepatitis C. Journal of Hepatology 69, 1088–1098 (2018).

[9] Shen, D., Sha, L., Yang, L. & Gu, X. Identification of multiple complications as independent risk factors associated with 1-, 3-, and 5-year mortality in hepatitis B-associated cirrhosis patients. BMC Infectious Diseases 25 (2025).

[10] Albert, A. et al. Review article: chronic hepatitis C natural history and cofactors. Alimentary Pharmacology and Therapeutics 22, 74–78 (2005).

[11] Chen, S. L. & Morgan, T. R. The natural history of hepatitis C virus (HCV) infection. International Journal of Medical Sciences 3, 47–52 (2006).

[12] Seeff, L. B. Natural history of chronic hepatitis C. Hepatology 36, S35–S46 (2002).

[13] Kawaguchi, Y. & Mizuta, T. Interaction between hepatitis C virus and metabolic factors. World Journal of Gastroenterology 20, 2888–2901 (2014).

[14] Ratziu, V., Heurtier, A., Bonyhay, L., Poynard, T. & Giral, P. An unexpected virus-host interaction–the hepatitis C virus-diabetes link. Alimentary Pharmacology & Therapeutics 22, 56–60 (2005).

[15] Hamdane, N. et al. HCV-induced epigenetic changes associated with liver cancer risk persist after sustained virologic response. Gastroenterology 156, 2313–2329 (2019).

[16] Jain, M. K. et al. Has access to hepatitis C virus therapy changed for patients with mental health or substance use disorders in the direct-acting-antiviral period? Hepatology 69, 51–63 (2019).

[17] Kuwelker, S. et al. High sustained virologic response rates, regardless of race or socioeconomic class, in patients treated with chronic hepatitis C in community practice using a specialized pharmacy team. Medicine (Baltimore) 102, e34183 (2023).

[18] Rauch, A., Kutalik, Z. et al. Genetic Variation in IL28B Is Associated With Chronic Hepatitis C and Treatment Failure: A Genome-Wide Association Study. Gastroenterology 138, 1338–1345 (2010).

[19] Ghouse, J. et al. Integrative common and rare variant analyses provide insights into the genetic architecture of liver cirrhosis. Nature Genetics 56 (2024).

[20] Trépo, E., Zucman-Rossi, J. & Nault, J.-C. Germline mutations and somatic mosaicism in steatotic liver diseases and related liver carcinogenesis. Nature Reviews Gastroenterology Hepatology (2026).

[21] Davis, G. L., Alter, M. J., El-Serag, H., Poynard, T. & Jennings, L. W. Aging of hepatitis C virus (HCV)-infected persons in the United States: a multiple cohort model of HCV prevalence and disease progression. Gastroenterology 138, 513–521 (2010).

[22] Konerman, M. A. et al. Machine learning models to predict disease progression among veterans with hepatitis C virus. PLoS One 14, e0208141 (2019).

[23] Lu, M. et al. Dynamic risk assessment for hepatocellular carcinoma in patients with chronic hepatitis C. Journal of Viral Hepatitis 30, 746–755 (2023).

[24] Lee, J. S. et al. Suboptimal performance of hepatocellular carcinoma prediction models in patients with hepatitis B virus-related cirrhosis. Diagnostics 13 (2023).

[25] Coppel, S. et al. Extra-hepatic comorbidity burden significantly increases 90-day mortality in patients with cirrhosis and high model for end-stage liver disease. BMC Gastroenterology 20, 302 (2020).

[26] Gupta, A. et al. Assessing the risk of further decompensation and survival in patients with cirrhosis with variceal bleeding as their first decompensation event. The American Journal of Gastroenterology 118, 833–839 (2023).

[27] Hu, Y. et al. Application of supervised and semisupervised learning prediction models to predict progression to cirrhosis in chronic hepatitis C. Artificial Intelligence in Health 2, 87–99 (2025).

[28] Minami, T. et al. Machine learning for individualized prediction of hepatocellular carcinoma development after the eradication of hepatitis C virus with antivirals. Journal of Hepatology 79, 1006–1014 (2023).

[29] Al Alawi, A. M., Al Shuaili, H. H., Al-Naamani, K., Al Naamani, Z. & Al-Busafi, S. A. A Machine Learning-Based Mortality Prediction Model for Patients with Chronic Hepatitis C Infection: An Exploratory Study. Journal of Clinical Medicine 13, 2939 (2024).

[30] All of Us Research Program Investigators. The “All of Us” Research Program. New England Journal of Medicine 381, 668–676 (2019).

[31] Kathiresan, N. et al. Representation of Race and Ethnicity in the Contemporary US Health Cohort All of Us Research Program. JAMA Cardiology 8, 859–864 (2023).

[32] Yang, H. et al. Multi-modality risk prediction of cardiovascular diseases for breast cancer cohort in the All of Us Research Program. Journal of the American Medical Informatics Association 31, 2800–2810 (2024).

[33] Hsu, W.-C., Crowley, A. & Parzynski, C. S. The impact of different censoring methods for analyzing survival using real-world data with linked mortality information: a simulation study. BMC Medical Research Methodology 24, 203 (2024).

[34] Raman, S. R. et al. Analyzing missingness patterns in real-world data using the SMDI toolkit: application to a linked EHR-claims pharmacoepidemiology study. BMC Medical Research Methodology 24, 246 (2024).

[35] Yang, X. et al. Predicting interval from diagnosis to delivery in preeclampsia using electronic health records. Nature Communications 16, 3496 (2025).

[36] Quistrebert, J. et al. Genome-wide association and gene-virus interaction study of liver disease in hepatitis C virus-infected patients. medRxiv (2025). https://www.medrxiv.org/content/early/2025/10/17/2025.10.12.25337816.

[37] Chen, Y. et al. Genome-wide association meta-analysis identifies 17 loci associated with nonalcoholic fatty liver disease. Nature Genetics 55, 1640–1650 (2023).

[38] Xiao, Q.-A. et al. Identification of novel drug targets for liver cirrhosis and its potential side-effects by human plasma proteome. Scientific Reports 14 (2024).

[39] De Vincentis, A. et al. A Polygenic Risk Score to Refine Risk Stratification and Prediction for Severe Liver Disease by Clinical Fibrosis Scores. Clinical Gastroenterology and Hepatology 20, 658–673 (2022).

[40] Salameh, H. et al. PNPLA3 polymorphism increases risk for and severity of chronic hepatitis C liver disease. World Journal of Hepatology 8, 1584–1592 (2016).

[41] Ursu, L. D. et al. Clinical and histopathological changes in different KIR gene profiles in chronic HCV Romanian patients. International Journal of Immunogenetics 48, 16–24 (2021).

[42] Su, C.-W. Towards a more comprehensive evaluation of liver fibrosis in patients with chronic hepatitis C. Journal of the Chinese Medical Association 82, 807–808 (2019).

[43] Taura, N. et al. Relationship of α-fetoprotein levels and development of hepatocellular carcinoma in hepatitis c patients with liver cirrhosis. Experimental and Therapeutic Medicine 4, 972–976 (2012).

[44] Calvaruso, V. et al. Liver and cardiovascular mortality after hepatitis C virus eradication by DAA: data from RESIST-HCV cohort. Journal of Viral Hepatitis 28, 1190– 1199 (2021).

[45] Ye, Q.-X. et al. Short-term prognostic factors for hepatitis B virus-related acute-on-chronic liver failure. World Journal of Clinical Cases 10, 8186–8198 (2022).

[46] Lescot, T., Karvellas, C., Beaussier, M., Magder, S. & Riou, B. Acquired Liver Injury in the Intensive Care Unit. Anesthesiology 117, 898–912 (2012).

[47] Xu, J.-H.Yu, Y.-Y. & Xu, X.-Y. Management of chronic liver diseases and cirrhosis: current status and future directions. Chinese Medical Journal 133, 2647–2649 (2020).

[48] Moon, A. M., Singal, A. G. & Tapper, E. B. Contemporary epidemiology of chronic liver disease and cirrhosis. Clinical Gastroenterology and Hepatology 18, 2650–2666 (2020).

[49] Chang, W. H., Mueller, S. H., Chung, S.-C., Foster, G. R. & Lai, A. G. Increased burden of cardiovascular disease in people with liver disease: unequal geographical variations, risk factors and excess years of life lost. Journal of Translational Medicine 20, 2 (2022).

[50] Cholankeril, G. et al. Longitudinal changes in fibrosis markers are associated with risk of cirrhosis and hepatocellular carcinoma in non-alcoholic fatty liver disease. Journal of Hepatology 78, 493–500 (2023).

[51] Burke, L. et al. Hepatocellular carcinoma risk scores for non-viral liver disease: A systematic review and meta-analysis. JHEP Reports 7, 101227 (2025).

[52] Johnson, P. J. et al. Evaluation of the aMAP score for hepatocellular carcinoma surveillance: a realistic opportunity to risk stratify. British Journal of Cancer 127, 1263–1269 (2022).

[53] Soucise, A. et al. Mortality Review Committee: Under-standing Inpatient and 30-Day Mortality at a Comprehensive Cancer Center. Journal of Palliative Medicine 27, 1598–1605 (2024).

[54] Ying, X. et al. Impact of Race and Neighborhood Socioeconomic Characteristics on Liver Cancer Diagnosis in Patients with Viral Hepatitis and Cirrhosis. Journal of Clinical and Experimental Hepatology 13, 568–575 (2023).

[55] Yi, S.-W., Choi, J.-S., Yi, J.-J., Lee, Y.-h. & Han, K. J. Risk factors for hepatocellular carcinoma by age, sex, and liver disorder status: a prospective cohort study in Korea. Cancer 124, 2748–2757 (2018).

